# Theta-Range SEEG Stimulation for Temporal Lobe Mapping: An Alternative to Conventional 1-Hz and 50-Hz Protocols

**DOI:** 10.64898/2026.03.31.26349175

**Authors:** Aube Darves-Bornoz, Emmanuel J. Barbeau, Marie Denuelle, Anne Calvel, Amaury De Barros, Zoé Darrasse, Kim Guines, Jean-Albert Lotterie, Luc Valton, Jonathan Curot

**Author notes:** **Corresponding Author:** Jonathan Curot, MD, PhD, Brain and Cognition Research Center, Pavillon Baudot, CHU Purpan, BP 25202, 31052 Toulouse Cedex, France.

## Abstract

**Objective:** Electrical brain stimulations (EBS) are central to epileptic network identification and functional mapping during stereo-electroencephalography (SEEG), yet stimulation frequencies remain empirical, and standardized across patients and brain regions, producing false negatives and false positives, and potentially compromising surgical outcome. We investigated theta-range EBS (7 Hz) in the temporal lobe, a prominent physiological frequency band in this region, and compared it with conventional 1-Hz and 50-Hz protocols.

**Methods:** We analyzed 1,408 temporal EBS in 25 patients with drug-resistant epilepsy. Epileptic responses (afterdischarges, seizures) and clinical signs were assessed across the epileptic network and temporal structures (amygdala, hippocampus, neocortex, parahippocampal gyrus, white matter), and analyzed according to stimulation parameters (frequency, intensity, duration, total charge).

**Results:** At matched intensity and duration, 7-Hz EBS were associated with a higher occurrence of afterdischarges and clinical signs than 1-Hz EBS in several temporal structures (e.g., parahippocampal epileptogenic zone: p=0.014). Effects on usual seizure induction were less consistent. Comparisons with 50 Hz showed no systematic significant differences, with responses observed at one or both frequencies depending on structure and outcome. When controlling for total charge, frequency-related differences were attenuated. Some effects were sporadically observed at both intermediate frequency and charge quantity. No adverse events occured.

**Significance:** Theta-range stimulation modulates electrophysiological and clinical responses during SEEG mapping and may provide complementary information to conventional frequencies. These findings support exploring a broader range of stimulation frequencies, rather than relying solely on standard protocols.

## Introduction

Why does stereo-electroencephalography still predominantly rely on only two stimulation frequencies—1 Hz and 50 Hz—in routine clinical mapping? This widespread practice remains largely empirical, with limited objective justification ^1,2^. Although epilepsy surgery can achieve seizure freedom in up to 70% of patients with drug-resistant epilepsy ^3^, pre-surgical evaluation using SEEG or electrocorticography is lengthy, often requiring one to three weeks ^4,5^. Several strategies—including interictal biomarkers such as fast ripples ^6^, virtual brains ^7^, and seizures induced by direct electrical brain stimulation (EBS) ^8^—aim to optimize this process by reducing monitoring duration and waiting time for spontaneous seizures, and improving presurgical evaluation and surgical outcomes.

However, EBS suffer from low reproducibility ^2^ and carry risks of false negatives (no effect in eloquent cortex ^9^) and false positives (afterdischarges in healthy tissue ^10^, unusual seizures ^11^). These limitations compromise the outcomes of pre-surgical evaluations and reflect an incomplete understanding of EBS mechanisms at circuit and network levels ^12^, as well as insufficient research on parameter settings. Current protocols rely on empirical electrical settings unchanged since the 1960s, despite SEEG being patient-specific ^1,4^. Similar empiricism persists across all fields of intracranial stimulation, including therapeutic neuromodulation ^13,14^.

In neuromodulation, highly variable outcomes motivate to enhance its predictability and effectiveness ^15,16^. Neuronal populations exhibit frequency-dependent resonance, with maximal responses often occurring near endogenous oscillations ^17^, and local networks can be entrained at the stimulation frequency ^18–20^. Recent work in deep brain stimulation (DBS) shows that frequency modulates functional connectivity non-linearly ^21^, highlighting the limitations of restricting SEEG stimulations to only “low” and “high” frequencies. Accordingly, intermediate frequencies have proven informative, such as 6–12 Hz EBS improving language mapping ^22^.

These observations support the hypothesis that tailoring SEEG stimulations to local physiological activity ^23^ may enhance mapping efficacy through frequency-specific network engagement. For example, applying EBS with theta-band frequency EBS in the temporal lobe, where networks frequently operate within this range, warrants consideration ^23,24^.

In this prospective study, we therefore evaluated a frequency more adapted to the physiological resting-state activity ^23^ of temporal structures—regions often involved in drug-refractory epilepsy. The frequency of 7 Hz was selected to reflect dominant oscillatory activity across temporal subregions. We compared 7-Hz EBS with standard 1-Hz and 50-Hz protocols for their ability to evoke epileptic effects (seizures, afterdischarges) and clinical signs (e.g., motor symptoms, memory phenomena…). To our knowledge, this is the first protocol readily applicable to clinical routine, that combines a prospective clinical comparison for both epileptic and functional mappings, controlling for frequency and other parameters (intensity, duration) during SEEG, with a frequency selection based on a rational, physiologically-grounded hypothesis, rather than arbitrary choices. By selecting a lobar-specific frequency, this readily applicable approach aims to improve diagnostic yield while capturing information typically obtained using both low- and high-frequency EBS.

## 2. Methods

### Patients and Stimulation Procedure

#### Patients

Twenty-five patients with drug-resistant focal epilepsy underwent invasive electrophysiological investigations at Toulouse University Hospital (November 2018–January 2024) to delineate the epileptogenic zone (EZ). Patients were implanted with SEEG intracranial depth electrodes, tailored to clinical requirements, with monitoring lasting 1–2 weeks (median ± SD: 11.0 ± 3 days), until sufficient seizures and clinical data were obtained. Antiseizure medications were reduced during monitoring. Demographic and clinical data were systematically collected.

Patients were enrolled in the StiMiC trial (NCT03738072), which allowed using additional EBS frequencies. They were selected based on an atlas of physiological intracranial resting-state oscillations ^23^, reflecting dominant frequency bands observed in each lobe’s subregions. One representative frequency per lobe was predefined accordingly.

Prior to participation, all patients provided written informed consent for the use of their data in research and for experimental EBS. The study was approved by the local Ethics Committee (CPP Ile de France VII; ID:18-026) and the French National Agency for Medicines and Health Products Safety (ANSM; 2018-A00383-52).

#### Electrode Implantation and Localization

Patients were implanted with standard depth electrodes and micro-macroelectrodes (Microdeep and Microdeep Micro-Macro; DIXI Medical, France) ^25,26^. Implantation planning (electrode location, direction, length) considered clinical hypotheses about epileptic networks, and anatomical or vascular constraints. Postoperatively, electrode were located by co-registering post-implantation CT scans with preoperative 3D T1-weighted MRI, allowing visual verification of contact locations.

Contacts were mapped into normalized MNI ICBM152 space using Brainstorm ^27^. Anatomical labels (AAL3 atlas ^28^) and gray/white matter classification were assigned according to the majority tissue composition within a 4-mm radius sphere centered on each contact pair (Fig. 1). Contact pairs were grouped into anatomical regions: amygdala, hippocampus, temporal neocortex, parahippocampal gyrus, and temporal white matter. White matter contacts stimulated were usually juxtacortical.

**Figure 1:**
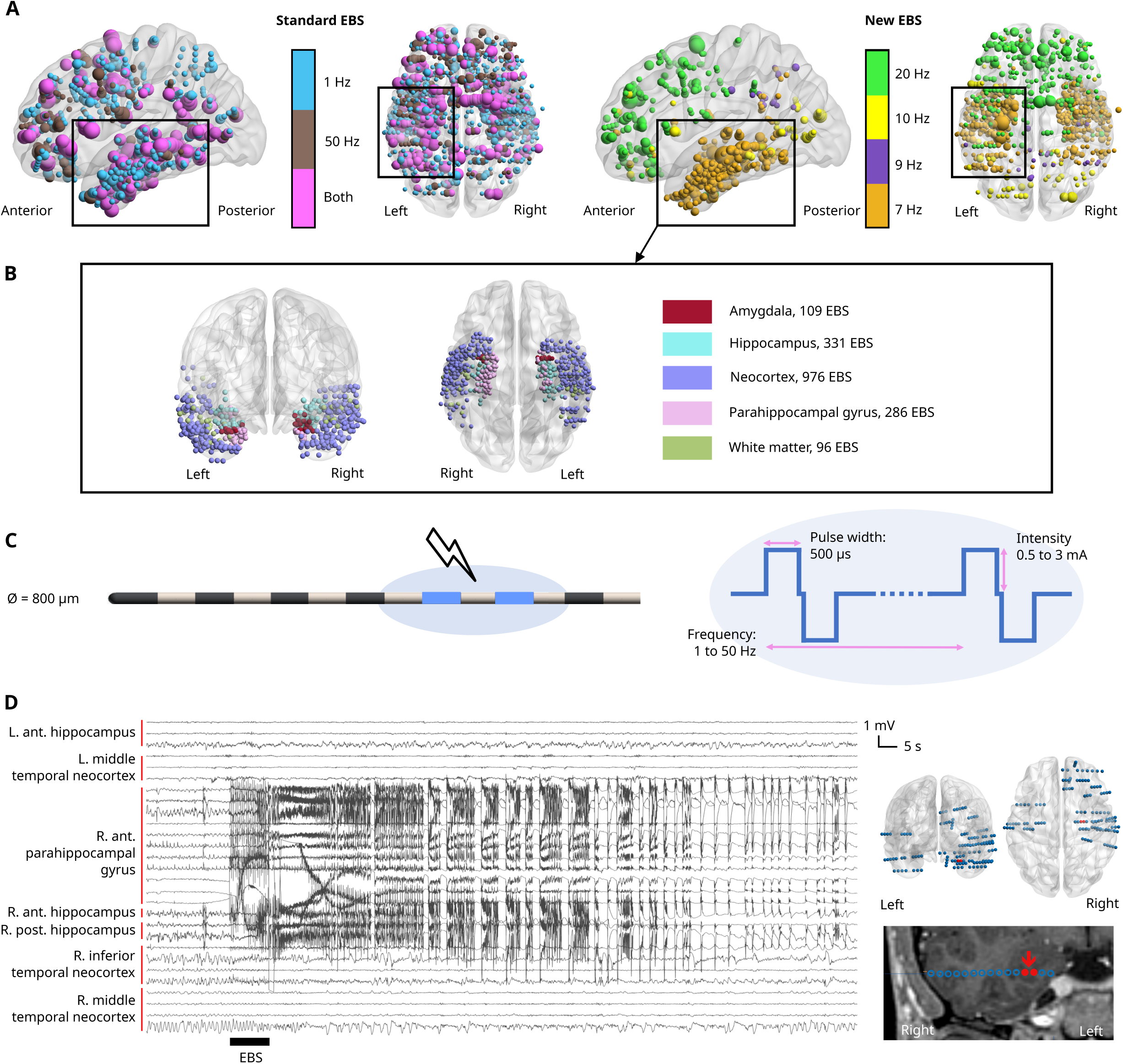
EBS dataset and procedure. (A) Location in normalized space of the contact pairs stimulated across all brain lobes (top) at each frequency studied (standard: top-left, experimental: top-right). The sphere diameter is proportional to the number of EBS applied. Colors represent the frequency used. (B) Location in normalized space of the contact pairs stimulated in each temporal structure studied. Colors represent the temporal structure. (C) Electrode and EBS configuration. (D) Example of an EBS in the right anterior parahippocampal gyrus, which induced an after-discharge, high-pass filtered at 0.1 Hz (left). Patient implantation (connected contacts only), with stimulated contact pair in red (top-right), and stimulated electrode on the MRI (bottom-right). The stimulated macrocontacts are indicated by the red arrow.

#### Recording Systems

SEEG was recorded continuously, using two SystemPLUS EVOLUTION 64-channel acquisition units (Micromed, Italy) at a sampling rate of 2048 Hz. A white matter macro-contact served as a reference.

#### Epileptic Network Definition

The lesion was defined on the MRI. The EZ was defined as the presumed regions from which seizures originate, and the irritative zone (IZ) as regions from which interictal activity is recorded ^29^. They were identified by the clinicians, who were blinded to the study outcomes (MD, LV). The EZ and IZ could overlap, therefore, additional analyses examined the IZ outside the EZ, to better assess epileptic events arising from regions outside the EZ. Non-involved cortex was classified as structures outside the lesion, EZ and IZ.

#### EBS Parameters

EBS sessions started after the first spontaneous seizure and were conducted by a clinician (MD, LV, JC). Each session lasted up to one hour or was stopped if a seizure or fatigue occurred. Two to seven sessions were conducted per patient, based on clinical needs.

EBS consisted of alternating biphasic square-wave 500-µs pulses delivered between contiguous contact pairs using a current generator (SD LTM Stimulator, Micromed).

Each contact pair was stimulated at either a standard frequency (1 Hz or 50 Hz) or at 7 Hz (Fig. 1). The clinician could start with either 1-Hz or 7-Hz EBS, with no imposed sequence order. To note, however, that the order of frequencies could not be fully randomized and was adapted to clinical constraints. To limit potential sequence effects, consecutive EBS on the same contact pair were avoided, and a minimum interval of 30 s separated successive EBS. This interval was extended when afterdischarges or seizures occurred, until return to pre-stimulation level. For paired comparisons, each 7-Hz EBS was applied at the same intensity and duration as a corresponding standard EBS on the same contact pair. Paired EBS were administered during the same EBS session, ensuring that the difference in effects observed was only caused by the choice of parameters and not by the level of antiseizure medications and elapsed time since implantation or last seizure. For example, if a 1-Hz EBS was performed at 2.5 mA for 10 s, a 7-Hz EBS was also applied at 2.5 mA for 10 s on the same contacts, in the same EBS session. Additional EBS were sometimes applied, with similar or different intensity or duration.

Intensity ranged from 0.5 to 3 mA, and the duration was 5, 10, or 20 s, following clinical hypotheses on tissue excitability and empirical clinical guidelines used in French epilepsy centers ^4^. Patients were blinded to stimulation timing, and sham trials were included, during which patients were asked if they had felt anything despite no EBS being applied.

Total charge quantity (µC) was calculated retrospectively for each EBS as the product of intensity, pulse duration, frequency, and total EBS duration.

#### Evaluation of EBS Effects

We focused on clinically relevant effects for defining the EZ and functional mapping: *epileptic effects* and *clinical signs*.

*Epileptic effects* were defined as all epileptic changes induced by EBS, including both afterdischarges and seizures. *Afterdischarges* were characterized as transient <u>electrographic</u> changes occurring immediately after EBS ^9,11,30^. *Seizure*s were defined as a sequence of clinical symptoms associated with concomitant electrographic discharges ^26^. These could either replicate the patient’s habitual semiology (“*usual seizures*”) or present atypical features (“*unusual seizures*”). Only *usual* seizures were included in the main analyses. *Non-usual* seizures were only reported descriptively.

*Clinical signs* encompassed any sign elicited by EBS, whether perceived by the patient or observed by the examiner, seizure-related or independent of seizure activity, associated with visible electrographic changes or not. Clinical signs provided critical information for both diagnostic and functional mapping, even when unrelated to seizure activity. Signs were classified according to the International League Against Epilepsy (ILAE) terminology ^31^: symptoms related to consciousness (e.g., impaired awareness), sensory perception, affect (e.g., fear), cognition (e.g., involuntary memories, language disturbances), autonomic functions (e.g., piloerection), and either elementary (e.g., eyelid clonus) or complex motor activities. *Adverse effects of EBS* included pain at the stimulation site, uncontrolled seizures/status epilepticus, or permanent neurological deficits attributable to EBS.

## Statistical Analyses

The objective was to isolate the effect of frequency on electrophysiological and clinical responses. Paired 7-Hz and standard EBS were compared on the same contact pair with identical intensity and duration, thereby minimizing confounding effects from EBS parameters, anatomy, or interindividual variability.

Because total charge quantity differs between two EBS delivered at different frequencies (for identical intensity and duration), we assessed the potential influence of charge quantity on outcomes. Comparisons at equal charge quantities were not feasible, as EBS were prioritized to match intensity and duration. Instead, EBS were grouped into charge quantity intervals (0–50, 50–100, 100–150, 150–200, 200–300, 300–1000 µC), and paired within these intervals.

Since multiple EBS on the same contacts were sometimes applied with identical parameters, responses were summarized as a proportion score (0: no effect in none of the EBS, up to 1: effect after each EBS) for the set of EBS matching each unique combination of {patient × contact pair × frequency × intensity × duration} or {patient × contact pair × frequency × charge interval}. Proportion scores were paired per frequency and compared using Wilcoxon signed-rank tests (1 Hz vs. 7 Hz or 7 Hz vs. 50 Hz).

EBS were excluded when no matching stimulation with identical parameters on the same contact pair had been delivered at the other frequency (mainly due to patient fatigue or seizure-related interruption).

Epileptic effects were analyzed across four epileptic network regions and five anatomical structures, yielding 20 combinations. Clinical signs (seven ILAE categories) were analyzed across the four grey-matter regions, yielding 28 conditions.

Statistical significance was set at p<0.05, with Benjamini–Hochberg false-discovery rate (FDR) correction applied within each effect type. Effect sizes were reported as absolute rank-biserial correlations |r_B_| (> 0.1: small, > 0.3: medium, > 0.5: large).

Sensitivity and specificity for inducing usual seizures were compared across frequencies using McNemar’s test. Sensitivity was defined as the proportion of stimulated contact pairs within the EZ that induced at least one usual seizure; specificity as the proportion of stimulated contact pairs outside the EZ that induced none.

Finally, in order to assess a potential effect of aetiology, responses were compared between patients with and without a mesial temporal EZ using Mann–Whitney U tests, with FDR correction applied within each effect category (e.g., percentage of afterdischarges in the hippocampal IZ at 1 Hz).

## 3. Results

### Population and Clinical Characteristics

Patients were implanted with 13.0 ± 1.6 electrodes. MRI revealed visible lesions in 23 patients, including 15 temporal lesions. Although all patients were explored in at least one temporal region, the EZ eventually involved the temporal lobe in 19 patients.

Eleven patients underwent resective surgery, with six achieving an Engel Class I outcome at one year; one additional patient was seizure-free at one-year follow-up after thermocoagulation; two patients were awaiting surgery. Surgery was contraindicated in nine patients. One of them was contraindicated but is waiting for a 2^nd^ SEEG for deep thermocoagulation (see individual clinical characteristics in Table 1). Two patients required a second SEEG evaluation.

**Table 1:**
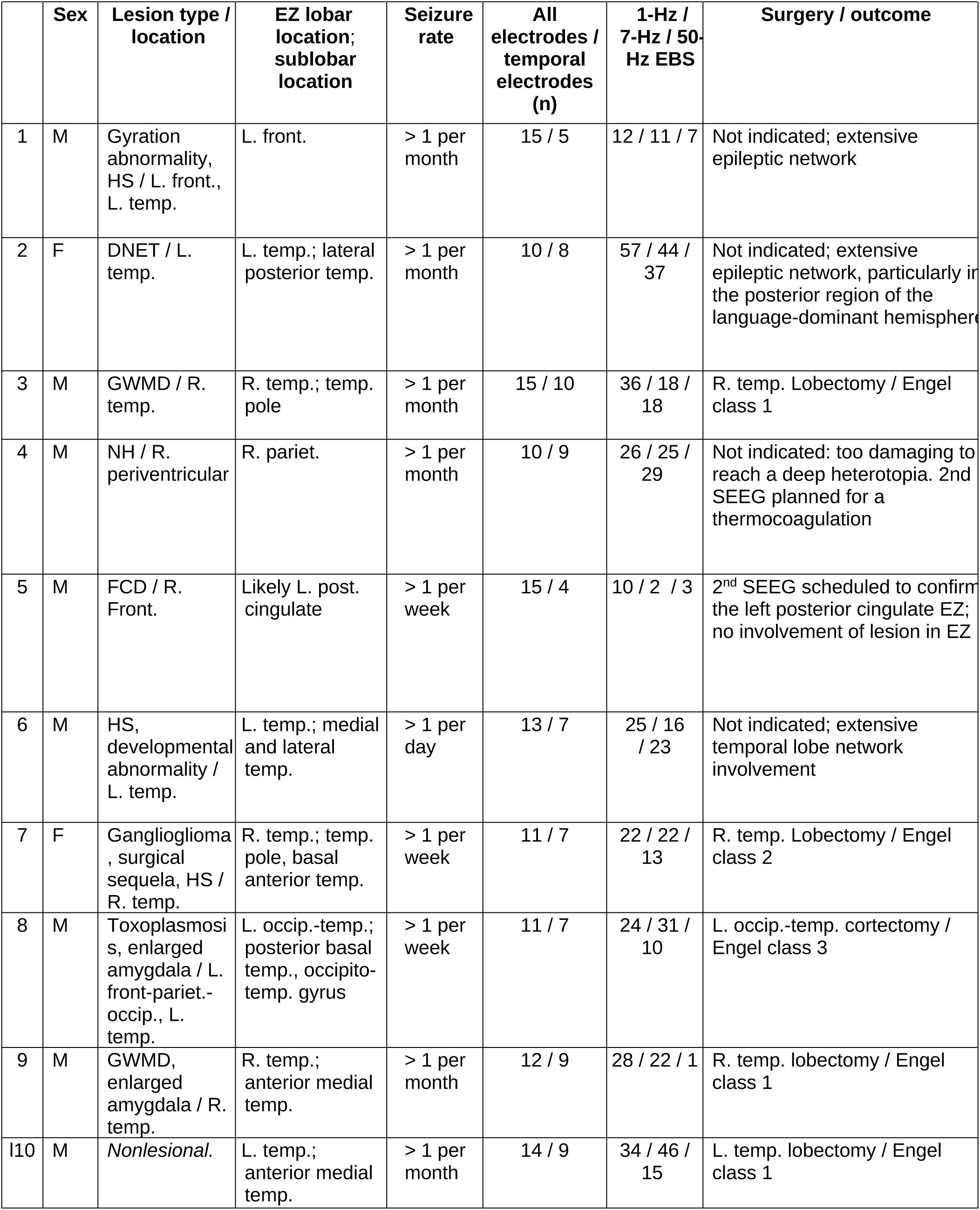
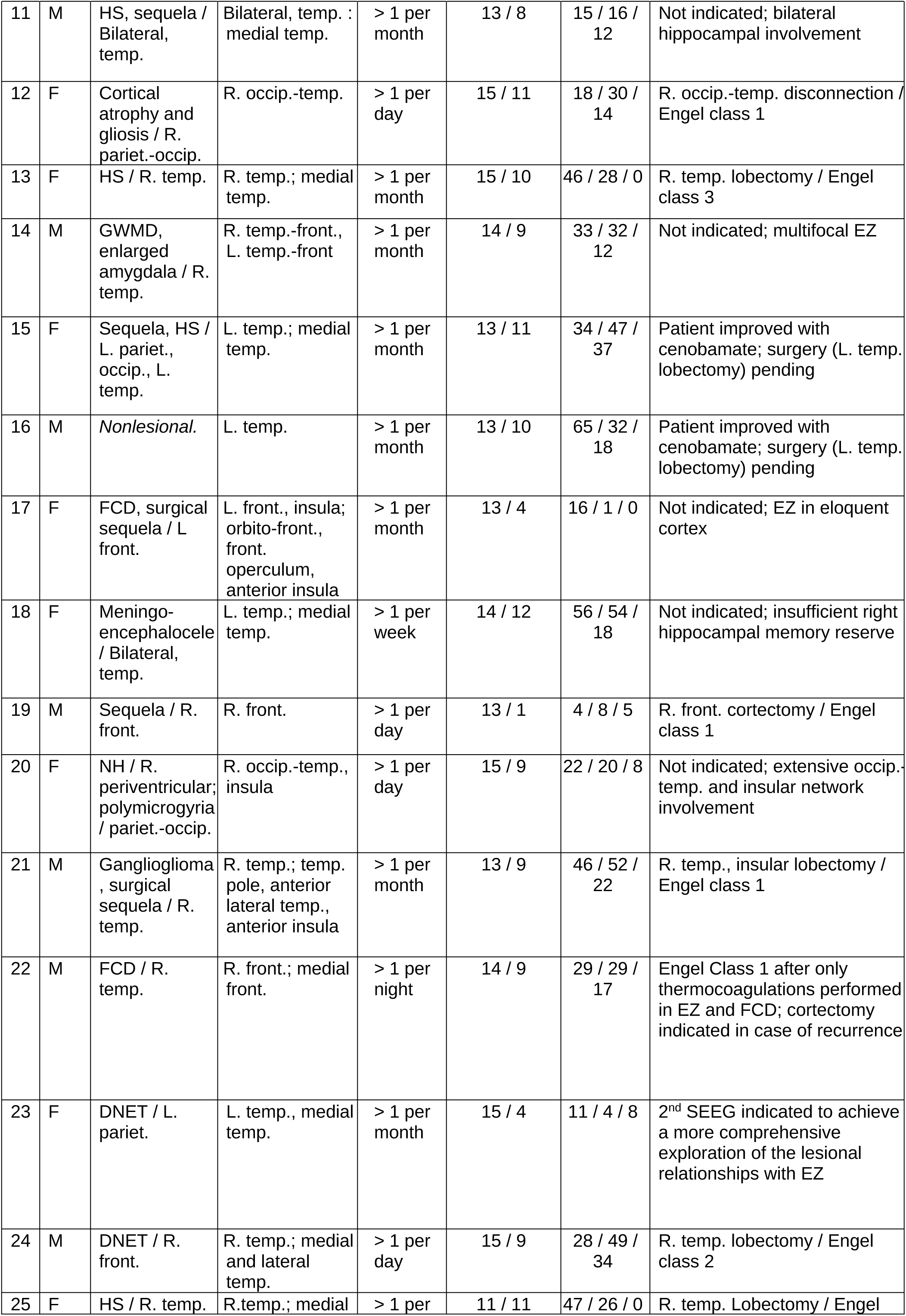
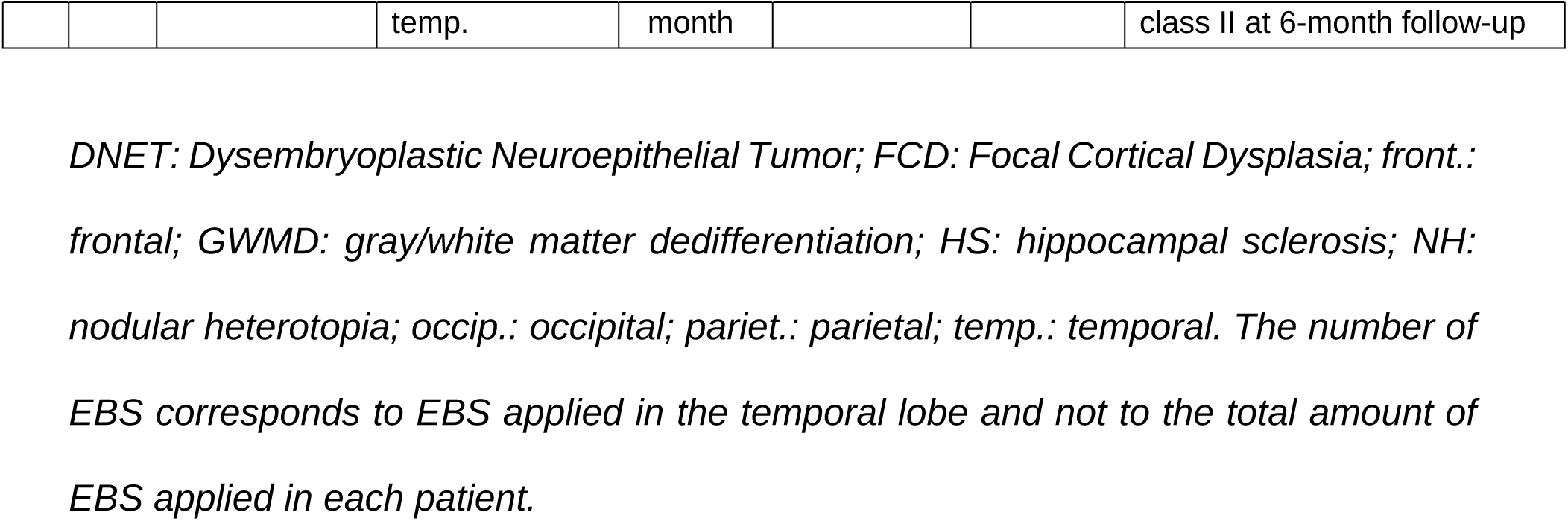
Clinical characteristics of patients stimulated in the temporal lobe.

1,810 temporal EBS were performed in 25 patients (median ± SD: 32.0 ± 11.8 years old; 15 men). Among these, 1,408 EBS could be paired and included: 578 at 1 Hz, 645 at 7 Hz, and 185 at 50 Hz. No adverse effects occurred. Paired comparisons at identical intensity and duration included 436 1-Hz versus 433 7-Hz EBS, and 159 50-Hz versus 154 7-Hz EBS, across all patients. Charge-matched comparisons were available for 248 1-Hz versus 222 7-Hz EBS, and for 29 50-Hz versus 28 7-Hz EBS across 21 patients (patients 6, 14, 17, 20 not included due to unavailable matches). The distribution of EBS effects across electrical parameters is shown in Fig. S1.

### Differential Afterdischarge Occurrence According to EBS Frequency

At identical intensity and duration, afterdischarges were more frequently observed during 7-Hz than 1-Hz EBS across multiple temporal structures, with large effect sizes. Within the EZ, a higher occurrence at 7 Hz compared with 1 Hz was found in the hippocampus (p=0.048, |r_B_|=0.788), neocortex (p=0.034, |r_B_|=1.000), and parahippocampal gyrus (p=0.014, |r_B_|=0.970) (Fig. 2; Table S2). In the amygdala and white matter, afterdischarges were observed only at 7 Hz, although these differences were not statistically significant.

**Figure 2:**
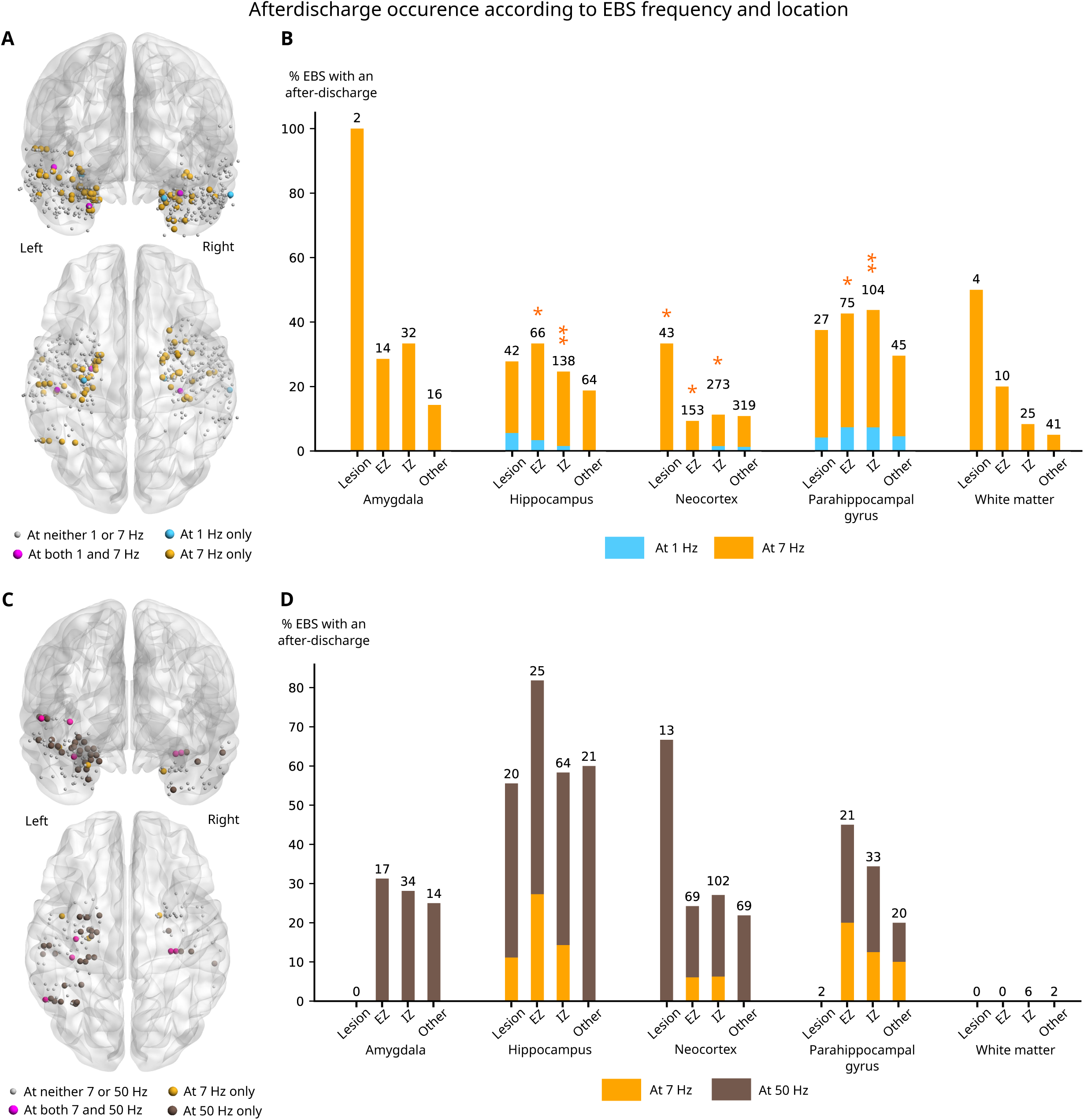
Proportion of afterdischarges induced by EBS, according to EBS frequency and location, compared at identical intensity and duration. *Afterdischarges* were characterized as transient electrographic changes occurring immediately after EBS. These events may remain localized to the stimulated area or propagate to distant brain regions, with or without observable clinical correlates. Responses were summarized as a proportion score (0: no effect, in none of the EBS; 1: afterdischarge after each EBS) for the set of EBS matching each unique combination of {patient × contact pair × frequency × intensity × duration}. Proportion scores were paired per frequency and compared using Wilcoxon signed-rank tests (1 Hz vs. 7 Hz or 7 Hz vs. 50 Hz). (A) Location of contact pairs included in paired comparisons between 1-Hz and 7-Hz EBS. (B) Percentage of 1-Hz EBS (blue) and 7-Hz EBS (orange) eliciting an afterdischarge across temporal structures and epileptic network regions. The number of EBS in each paired comparison is noted above each column. (C) Location of contact pairs included in paired comparisons between 7-Hz and 50-Hz EBS. (D) Percentage of 7-Hz EBS (orange) and 50-Hz EBS (brown) eliciting an afterdischarge across temporal structures and epileptic network regions. Asterisks indicate significance based on Wilcoxon signed-rank tests after FDR correction for multiple comparisons (p<0.05: *; p<0.01: **).

A similar distribution was observed in the IZ, with a higher occurrence of afterdischarges at 7 Hz in the hippocampus (p=0.008, |r_B_|=0.896), neocortex (p=0.014, |r_B_|=0.846), and parahippocampal gyrus (p=0.008, |r_B_|=0.860). In the amygdala and white matter, afterdischarges were again observed only at 7 Hz, without reaching statistical significance. In the IZ outside the EZ, afterdischarges were more frequently observed at 7 Hz in the hippocampus (p=0.045, |r_B_|=1.000). In neocortical lesions, afterdischarges were observed only at 7 Hz (p=0.034, |r_B_|=1.000).

In contrast, no significant difference emerged between 7-Hz and 50-Hz EBS in any structure. When afterdischarges were observed (in 15 structures), they occurred either at both frequencies or exclusively at 50 Hz, but not significantly in any of the conditions.

Several afterdischarges were recorded in non-involved temporal regions at all frequencies studied, without a significant effect of EBS frequency.

When comparisons were performed with matched charge quantity, no statistically significant differences were detected between frequencies (Table S3). A higher occurrence at 7 Hz compared with 1 Hz was noted in the neocortical IZ (p_raw_=0.025, |r_B_|=0.929), a pattern that was already observed at equal intensity and duration, although this did not remain significant after FDR correction (p_FDR_=0.624).

### Differential Usual Seizure Occurrence According to EBS Frequency

Frequency-related differences in usual seizure induction were less marked than those observed for afterdischarges. Across temporal structures, usual seizures were observed exclusively (12 structures) or more frequently (six structures) during 7-Hz EBS compared with 1-Hz EBS, without reaching statistical significance (Fig. 3; Table S2). In contrast, when comparing 7-Hz to 50-Hz EBS, usual seizures were observed exclusively during 50-Hz stimulation in 11 structures, but again without significance.

**Figure 3:**
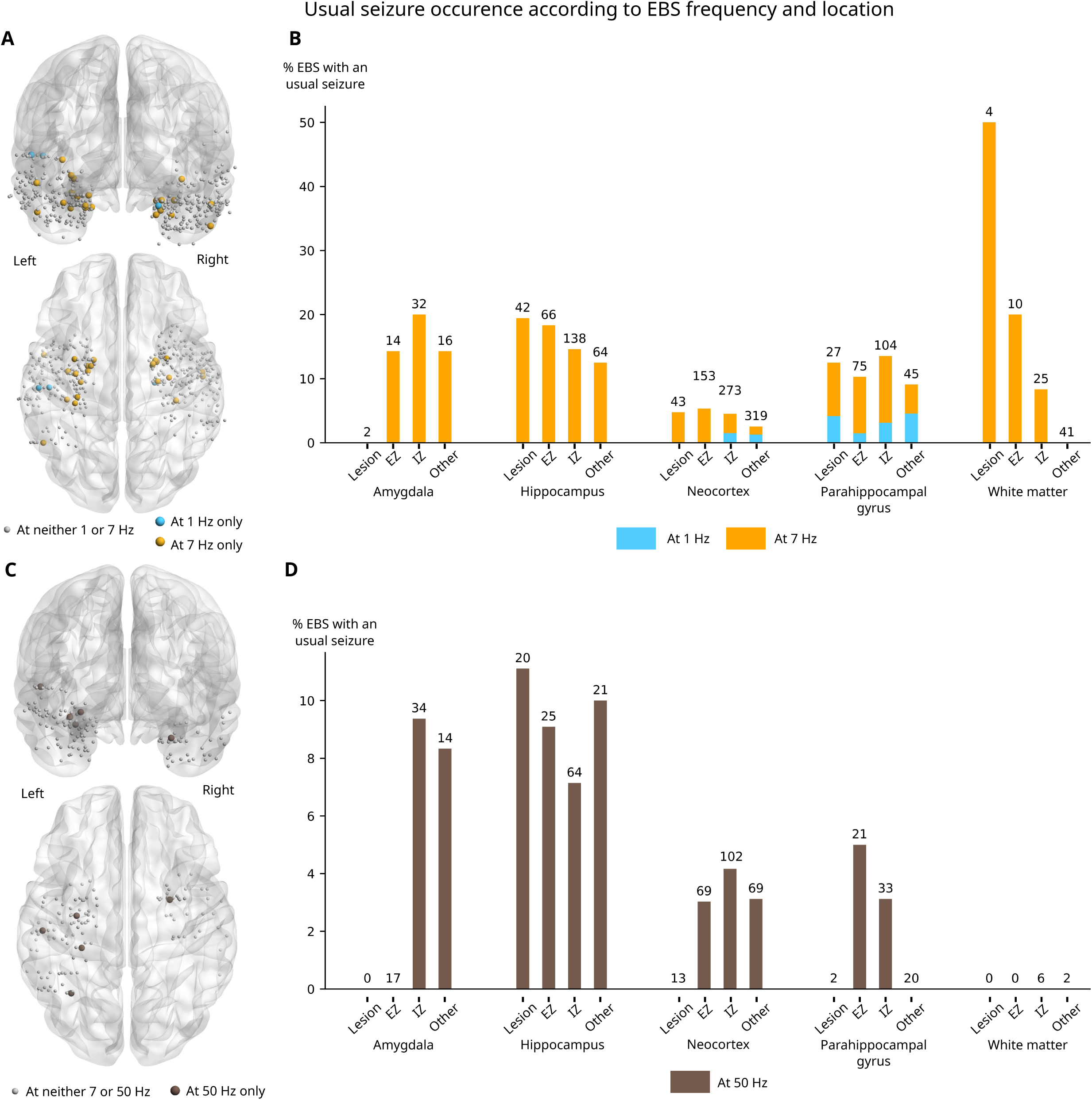
Proportion of usual seizures induced by EBS, according to EBS frequency and location, compared at identical intensity and duration. Responses were summarized as a proportion score (0: no effect, in none of the EBS; 1: usual seizure after each EBS) for the set of EBS matching each unique combination of {patient × contact pair × frequency × intensity × duration}. Proportion scores were paired per frequency and compared using Wilcoxon signed-rank tests (1 Hz vs. 7 Hz or 7 Hz vs. 50 Hz). (A) Location of contact pairs included in paired comparisons between 1-Hz and 7-Hz EBS. (B) Percentage of 1-Hz EBS (blue) and 7-Hz EBS (orange) eliciting a usual seizure across temporal structures and epileptic network regions. The number of EBS in each paired comparison is noted above each column. (C) Location of contact pairs included in paired comparisons between 7-Hz and 50-Hz EBS. (D) Percentage of 7-Hz EBS (orange) and 50-Hz EBS (brown) eliciting a usual seizure across temporal structures and epileptic network regions.

When pooling all temporal regions, sensitivity for inducing usual seizures from the EZ was significantly higher with 7-Hz than with 1-Hz EBS (9.6% vs 0.6%, p<0.001), while specificity did not differ significantly (98.9% vs 96.3%, p=0.09). No significant differences in sensitivity or specificity were observed between 7-Hz and 50-Hz EBS (0.0% vs 4.8%, p=0.25; 100.0% vs 95.3%, p=0.125).

When accounting for charge quantity, no statistically significant differences were identified across structures, although trending toward more usual seizures at 7 Hz when compared to 50 Hz (Table S3).

Sensitivity and specificity did not differ significantly when comparing 7-Hz EBS with 1-Hz EBS (3.1% vs 7.8%, p=0.25; 98.8% vs 97.6%, p=1.0) or with 50-Hz EBS (9.1% vs 0.0%, p=0.25; 76.9% vs 92.3%, p=0.50).

Two unusual seizures were recorded in patient 20 during 7-Hz EBS (fusiform gyrus, 2 mA, 10 s; hippocampus, 2.5 mA, 10 s; both IZ) and two in patient 10 during 50-Hz EBS (amygdala, 1.2 mA, 5 s, both IZ).

### Differential Clinical Sign Occurrence According to EBS Frequency

We compared the efficacy of 7-Hz EBS to induce clinical signs across 28 conditions (seven clinical categories across four temporal regions).

At identical intensity and duration, clinical signs were observed exclusively (12 conditions) or predominantly (12 conditions) during 7-Hz EBS compared with 1-Hz EBS. Statistically significant differences were identified for cognitive signs in the hippocampus (p=0.011, |r_B_|=1.000) and neocortex (p=0.011, |r_B_|=1.000), autonomic signs in the hippocampus (p=0.027, |r_B_|=0.818), and sensory signs in the neocortex (p=0.004, |r_B_|=0.778) and parahippocampal gyrus (p=0.005, |r_B_|=1.000) (Fig. 4; Table S4). In seven other conditions, differences did not reach significance, and proportions were similar between frequencies in two conditions. No condition showed a higher occurrence at 1 Hz.

**Figure 4:**
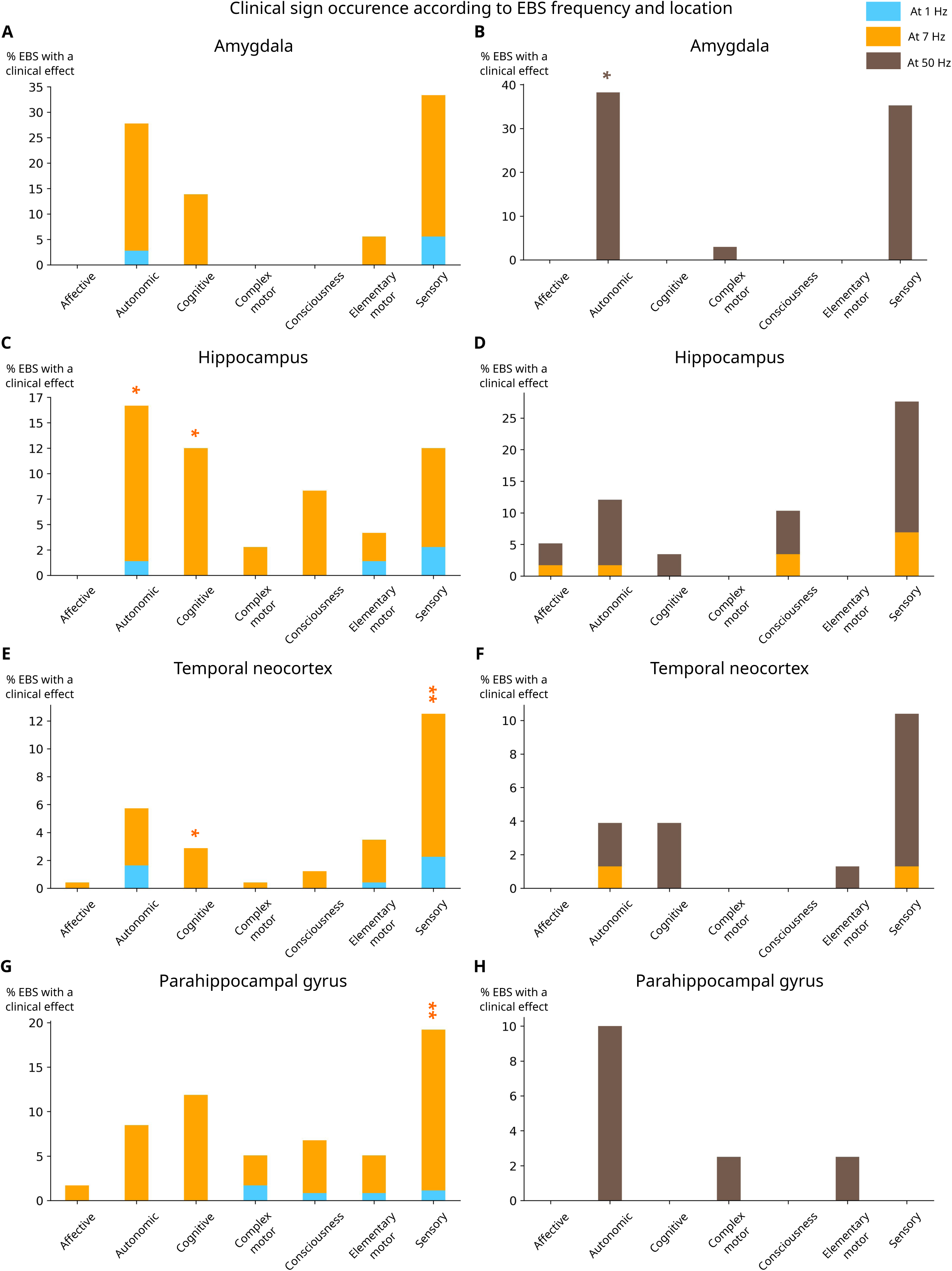
Proportion of clinical signs induced by EBS, according to EBS frequency and location, compared at identical intensity and duration. Signs were classified according to the International League Against Epilepsy (ILAE) terminology ^30^: symptoms related to consciousness (e.g., impaired awareness), sensory perception, affect (e.g., fear), cognition (e.g., involuntary memories, language disturbances), autonomic functions (e.g., tachycardia, piloerection), and either elementary (e.g., facial twitching, eyelid clonus) or complex motor activities. Responses were summarized as a proportion score (0: no effect, in none of the EBS; 1: clinical sign after each EBS) for the set of EBS matching each unique combination of {patient × contact pair × frequency × intensity × duration}. Proportion scores were paired per frequency and compared using Wilcoxon signed-rank tests (1 Hz vs. 7 Hz or 7 Hz vs. 50 Hz). For each temporal region, paired panels display comparisons of 1-Hz (blue) versus 7-Hz (orange) EBS and 7-Hz (orange) versus 50-Hz (brown) EBS: (A–B) amygdala (38 EBS), (C–D) hippocampus (152 EBS), (E–F) temporal neocortex (497 EBS), and (G–H) parahippocampal gyrus (127 EBS). Asterisks indicate significance based on Wilcoxon signed-rank tests after FDR correction for multiple comparisons (p<0.05: *; p<0.01: **).

Comparisons between 7 Hz and 50 Hz showed a more heterogeneous distribution. Clinical signs were observed in 15 conditions: occurring exclusively at 50 Hz in nine conditions—significantly for autonomic signs in the amygdala (p=0.046, |r_B_|=1.000). In the remaining six conditions, no statistically significant differences were identified.

When analyses were performed at matched charge quantity, no statistically significant differences in clinical sign occurrence were observed between frequencies, although a higher occurrence at 7 Hz was noted in some comparisons (Table S5).

An example of a cognitive sign elicited only at 7 Hz—at both intermediate frequency and charge quantity—is shown in Fig. 5.

**Figure 5:**
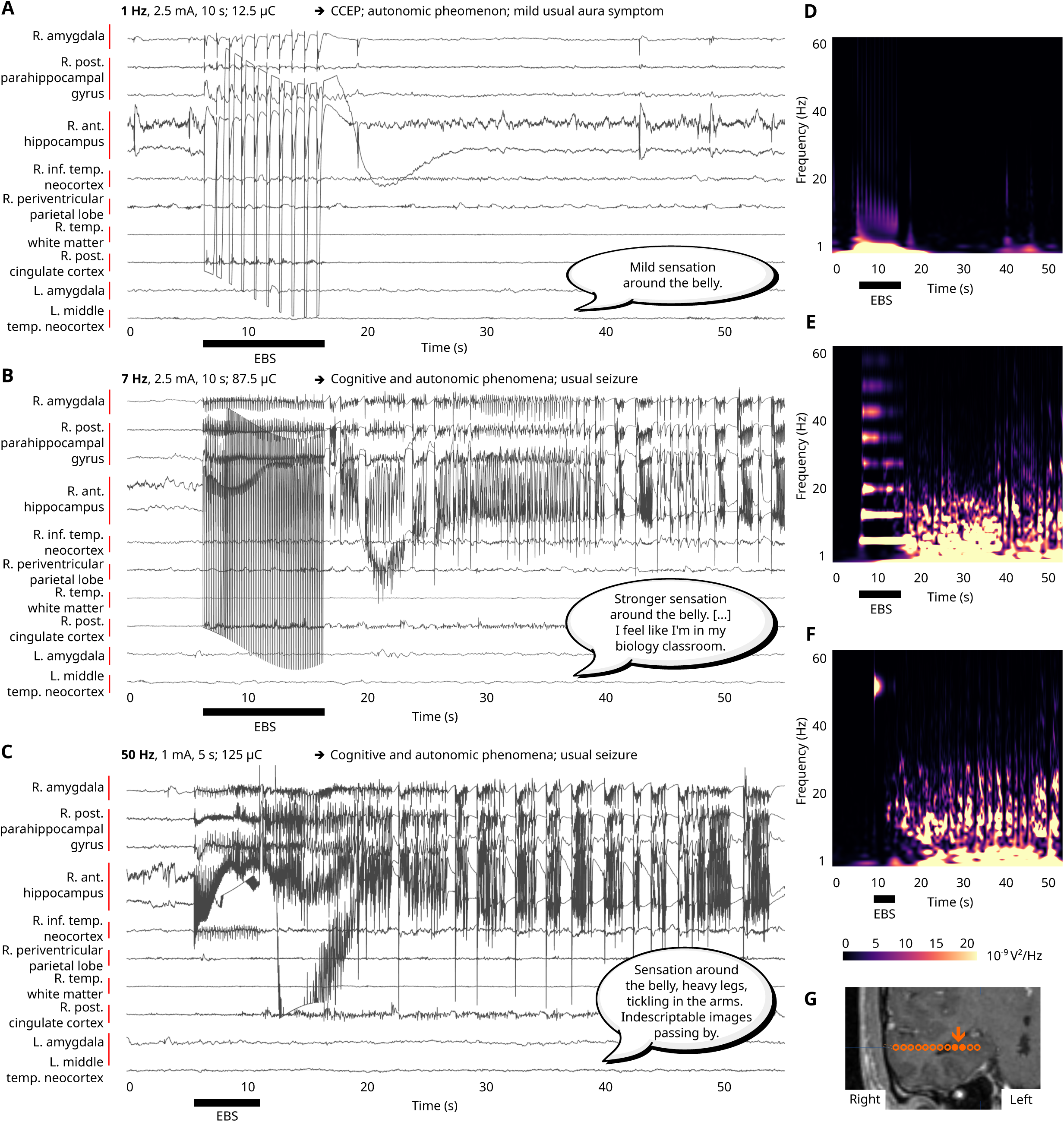
Comparison of EBS delivered on the same pair of contacts in the right anterior hippocampal IZ of Patient 4 (right temporal periventricular nodular heterotopia). In this patient, a 7-Hz EBS activated memory-related networks in the anterior hippocampus—a response absent with paired 1-Hz EBS and not clearly elicited under 50-Hz EBS. Panels show SEEG recordings at 1 Hz (A), 7 Hz (B), and 50 Hz (C). Corresponding time–frequency maps from an adjacent contact pair are shown for EBS at 1 Hz (D), 7 Hz (E), and 50 Hz (F). The anatomical location of the stimulated contact pair is displayed on MRI (G). At 1 Hz, a minor effect was observed (a mild vegetative aura symptom, no afterdischarge), while more intense effects were triggered at 7 Hz (an experiential phenomenon, with the induction of a reminiscence corresponding to a personal semantic memory, and an afterdischarge spreading within medial temporal structures). The 50-Hz EBS elicited an afterdischarge and a similar vegetative aura, but no experiential phenomenon was observed. EBS parameters, epileptic electrophysiological effects, and associated clinical signs are indicated in the corresponding panels, at 1 Hz (A), 7 Hz (B), and 50 Hz (C), with verbatim transcriptions. L: left; R: right.

Individual-level data are provided in Fig. S2–S4.

No significant differences were observed between patients with and without a temporo-mesial EZ for any clinical outcome (Tables S6–S8).

## 4. Discussion

Results indicate that EBS frequency influences electrophysiological and clinical responses during SEEG mapping. At matched intensity and duration, 7-Hz EBS was associated with a higher occurrence of afterdischarges and clinical signs than 1-Hz EBS across several temporal structures, whereas effects on usual seizure induction were less consistent, although pooled analyses showed higher sensitivity for EZ identification at 7 Hz without differences in specificity. In contrast, comparisons between 7-Hz and 50-Hz EBS did not reveal systematic differences across outcomes, with patterns depending on structure and measure. When controlling for total charge, frequency-related differences were attenuated and no longer significant. However, some responses were observed at both intermediate frequency and charge.

Taken together, these findings suggest that theta-range stimulation engages electrophysiological and clinical responses at least as effectively as conventional frequencies in several contexts, while revealing partially distinct response profiles. These results support broadening the range of stimulation frequencies explored during SEEG.

### Frequency-Dependent Effects and Implications for SEEG Mapping

Across temporal substructures, 7-Hz EBS were more frequently associated with afterdischarges and clinical signs than 1-Hz EBS, although differences were less consistent regarding usual seizures. This suggests that intermediate frequencies may engage epileptic and functional networks differently from low-frequency EBS, while maintaining a comparable safety profile.

In contrast, differences between 7-Hz and 50-Hz EBS were not consistent across structures or outcomes, and no systematic superiority emerged. However, 7-Hz EBS occasionally elicited more specific clinical signs than both standard frequencies (e.g., Fig. 5), suggesting a partial dissociation between seizure induction and functional mapping sensitivity, and frequency-dependent recruitment of network components depending on the context.

At the whole-temporal-lobe level, 7-Hz EBS were associated with higher sensitivity for inducing usual seizures from the EZ compared with 1 Hz, without loss of specificity, although this effect was not consistent across substructures and should be interpreted cautiously given uncertainties in EZ definition inherent to SEEG studies.

Overall, these findings support incorporating intermediate frequencies such as 7 Hz as complementary probes, particularly when responses are not elicited at standard frequencies or where additional functional characterization is required. While 1-Hz EBS remain essential for connectivity analyses, intermediate frequencies may complement 1-Hz protocols by increasing electrophysiological and clinical response yield, either as second-intention probes of excitability, alternatives to high-frequency EBS in selected conditions, or additional tests for functional probing in specific contacts.

In practice, this stepwise approach was reflected in our dataset. As in routine SEEG exploration, 1-Hz EBS were applied broadly, and 7-Hz EBS frequently performed alongside, whereas 50-Hz EBS were typically restricted to selected contacts, particularly when responses were absent or limited at lower frequencies or when additional probing was clinically indicated. This conditional application may have influenced response distributions, as 50-Hz EBS and their matched 7-Hz EBS were preferentially applied to less responsive sites. Conversely, the broader sampling of 7-Hz EBS when matched with 1 Hz may better reflect 7-Hz performance as an intermediate probe of network excitability.

Taken together, these observations support a stepwise stimulation strategy, combining low- and intermediate-frequency protocols, with more selective use of high-frequency stimulation, and provide a rationale for exploring a broader range of frequencies adapted to local network properties.

### Frequency is Not a Sole Proxy For Charge Quantity

When analyses were performed at comparable charge, frequency-related differences were attenuated and no longer statistically significant, indicating a substantial contribution of charge quantity to tissue excitability. This is consistent with prior work showing that similar behavioral or neural responses can be obtained with different combinations of stimulation parameters yielding equivalent charge ^32^.

However, this does not fully account for the persistence of frequency-dependent effects. In particular, trends remained in neocortical IZ, where afterdischarges were more frequently observed with 7-Hz than at 1-Hz EBS across both intensity/duration-and charge-matched conditions. Additionnally, certain responses occurred exclusively at 7 Hz and at intermediate charge, suggesting that frequency contributes beyond total charge alone. These findings align with evidence that stimulation effects depend on multidimensional interactions between frequency, intensity, and temporal dynamics ^33^, and may vary according to network state and localization ^34^.

Overall, these findings support the contribution of both charge-related and frequency-specific mechanisms to EBS effects, with their relative influence varying across structures and clinical contexts, consistent with an interaction between stimulation frequency and intrinsic network properties rather than a sole effect of delivered charge.

### Aligning EBS Frequency to Brain Rhythms

Synchronizing EBS frequencies with physiological oscillations may enhance neural engagement through local entrainment, improving epileptic and functional mapping ^35,36^. Neural resonance has been proposed as a biomarker of the EZ, with resonant frequencies inducing seizures and predicting surgical outcome ^37^, and the frequency-dependent patterns observed here are consistent with partial dependence of EBS effects on alignment with endogenous dynamics.

By potentially engaging theta-range activity, 7-Hz EBS may facilitate recruitment of temporal-lobe networks involved in seizure generation and functional responses, although this interpretation remains tentative and requires confirmation with combined spectral and connectivity analyses.

### Toward Personalized Theta-Frequency Stimulation

Although effective here, 7 Hz is not a universal optimal frequency for temporal-lobe mapping. Theta-band preferences likely vary across temporal cytoarchitectures and connectivity, reflecting physiological gradients, from low-theta (≈2–5 Hz) in the anterior hippocampus ^38,39^ to higher-theta (≈6–9 Hz) in posterior medial temporal regions, and neocortical theta–alpha patterns ^40^. The heterogeneous distribution of afterdischarges observed further supports substructure-specific frequency sensitivity, consistent with theta-burst EBS studies showing differential engagement of anterior and posterior hippocampal networks ^41^.

Theta-aligned EBS likely operates through both local resonance and network-level synchronization, as regions preferentially engage specific circuits at certain frequencies. For example, VNS and tDCS effects correlate with changes in long-range theta/alpha connectivity ^42,43^. Frequency-tuning approaches, exploring frequencies within targeted physiological windows, may therefore yield greater mapping specificity.

Optimal frequencies may also be individual-specific, as shown in Parkinson’s disease DBS ^44^ and memory-modulation studies ^45^, highlighting the potential of closed-loop stimulation, adjusting frequency to individual oscillations ^18,46^.

Future studies should explore broader and patient-specific frequencies, integrate seizure-onset patterns ^47^ to refine seizure induction and network mappings, and account for clinical factors like medication or fatigue ^35,36,44^ This highlights the need for multicenter studies combining systematic frequency exploration with physiological recordings and computational modeling. Such approaches may ultimately move SEEG stimulation from empiricism toward physiologically-informed, patient-specific neuromodulation.

## Conclusion

In a field where the clinical impact of EBS parameters remains incompletely defined, this study indicates that theta-range stimulation modulates electrophysiological and clinical responses during SEEG-based mapping in temporal lobe epilepsy. Compared with 1-Hz EBS, 7-Hz EBS was associated with a higher occurrence of afterdischarges and clinical signs in several contexts, while no systematic superiority was observed relative to 50-Hz EBS, but with some specific effects induced only at intermediate frequency and charge quantity. Theta-range EBS can provide complementary information, either as an alternative to high-frequency EBS in selected conditions or as an additional test for specific contacts.These findings suggest that intermediate frequencies engage epileptic and functional networks differently from conventional protocols, and support broadening the range of frequencies used during SEEG.

Although focused on temporal regions, these findings support the rationale for frequency tuning in SEEG, including in other regions. Future work should explore broader and patient-specific frequency ranges, including multiple theta-band values adapted to local substructures, and further refine comparisons with higher-frequency protocols to optimize EZ delineation.

Given the multidimensional nature of EBS, large collaborative datasets and combined with physiological recordings, computational modeling, and adaptive stimulation approaches are essential to better characterize frequency-dependent effects and to move toward more individualized, physiology-informed neuromodulation strategies in epilepsy ^48^.

## Data Availability

All data produced in the present study are available upon reasonable request to the authors

## Acknowledgments

We would like to thank the patients and nurses at the Neurology Department of Toulouse University Hospital, Aline Meulle, Adeline Gallini for administrative processes and methodological support to obtain funding, Simona Celebrini for her help in data acquisition, Ylias Kriket for data acquisition and selection, as well as Pierre Mégevand, Philippe Kahane, Stanislas Lagarde and Lionel Nowak for their valuable comments and inspiration. We also thank the LAAS Team, especially Ali Maziz, Elodie Despouy, Adrien Causse, Alexis Robin, Laure Balguerie, Hélène Mirabel, Jean-Christophe Sol, and the neuroradiology team.

The English syntax of some parts of this manuscript was corrected with the assistance of AI language models (ChatGPT; Grammarly) After using these tools, the content was reviewed and edited as needed, and all authors take full responsibility for the content of the manuscript. A native English speaker also corrected the syntax.

## Funding

This work was financially supported by the National Research Agency as part of the “DYNEUMICS” project, reference ANR-21-CE17-0029, and by Toulouse University Hospital (local grant: ARI StiMiC, NCT03738072).

## Author Contributions

AD-B, EB, JC contributed to the conception and design of the study. AD-B, ADB, J-AL, JC, LV, MD, ZD contributed to data acquisition. AC, AD-B, EB, JC, KG, LV, MD contributed to data analysis and interpretation. AD-B, EB, JC contributed to drafting the text and preparing the figures.

## Potential conflicts of Interest

None to report.

## Data Availability

The data that support the findings of this study are available on request from the corresponding author. The data are not publicly available due to privacy or ethical restrictions.

## Figure Legends

**Supplementary Figure S1:** Panel of electrical parameters (frequency, charge density, and duration) used in all EBS studied, and proportion of EBS that induced (A) an afterdischarge or (B) a clinical sign (whether related to epilepsy or not) for each of these electrical parameter combinations. The charge density of a single pulse was calculated from the product of intensity with the duration of the pulse (500 µs), divided by the surface of a macro-contact (0.05 cm²). Paired comparisons at identical intensity and duration were computed between 1-Hz and 7-Hz EBS at 2.0 ± 0.5 mA, 10.0 ± 1.8 s, 15.0 ± 31.6 µC, and between 50-Hz and 7-Hz EBS at 1.0 ± 0.4 mA, 5.0 ± 1.1 s, 70.0 ± 85.2 µC. Paired comparisons at matched charge quantities were computed between 1-Hz and 7-Hz EBS at 1.0 ± 0.7 mA, 10.0 ± 5.0 s, 15.0 ± 11.4 µC, and between 50-Hz and 7-Hz EBS at 1.0 ± 0.9 mA, 5.0 ± 2.5 s, 63.0 ± 22.6 µC.

**Supplementary Figure S2:** Percentage of afterdischarges and usual seizures induced by EBS per patient, according to EBS frequency and temporal structure, at identical intensity and duration. Panels show the proportions of 1-Hz (blue), 7-Hz (orange), and 50-Hz (brown) EBS eliciting afterdischarges or usual seizures. Temporal regions are displayed as follows: amygdala (A, E), hippocampus (B, F), temporal neocortex (C, G), and parahippocampal gyrus (D, H).

**Supplementary Figure S3:** Percentage of afterdischarges and usual seizures induced by EBS per patient, according to EBS frequency and epileptic network regions at identical intensity and duration. Panels show the proportions of 1-Hz (blue), 7-Hz (orange), and 50-Hz (brown) EBS eliciting afterdischarges or usual seizures. Epileptic network regions are displayed as follows: lesion (A, E), EZ (B, F), IZ (C, G), and non-involved (D, H).

**Supplementary Figure S4:** Percentage of clinical signs induced by EBS per patient, according to EBS frequency and temporal structure, at identical intensity and duration. Panels show the proportions of 1-Hz (blue), 7-Hz (orange), and 50-Hz (brown) EBS eliciting clinical signs. Temporal regions are displayed as follows: amygdala (A), hippocampus (B), temporal neocortex (C), and parahippocampal gyrus (D).

---

**Supplementary Table S1:**
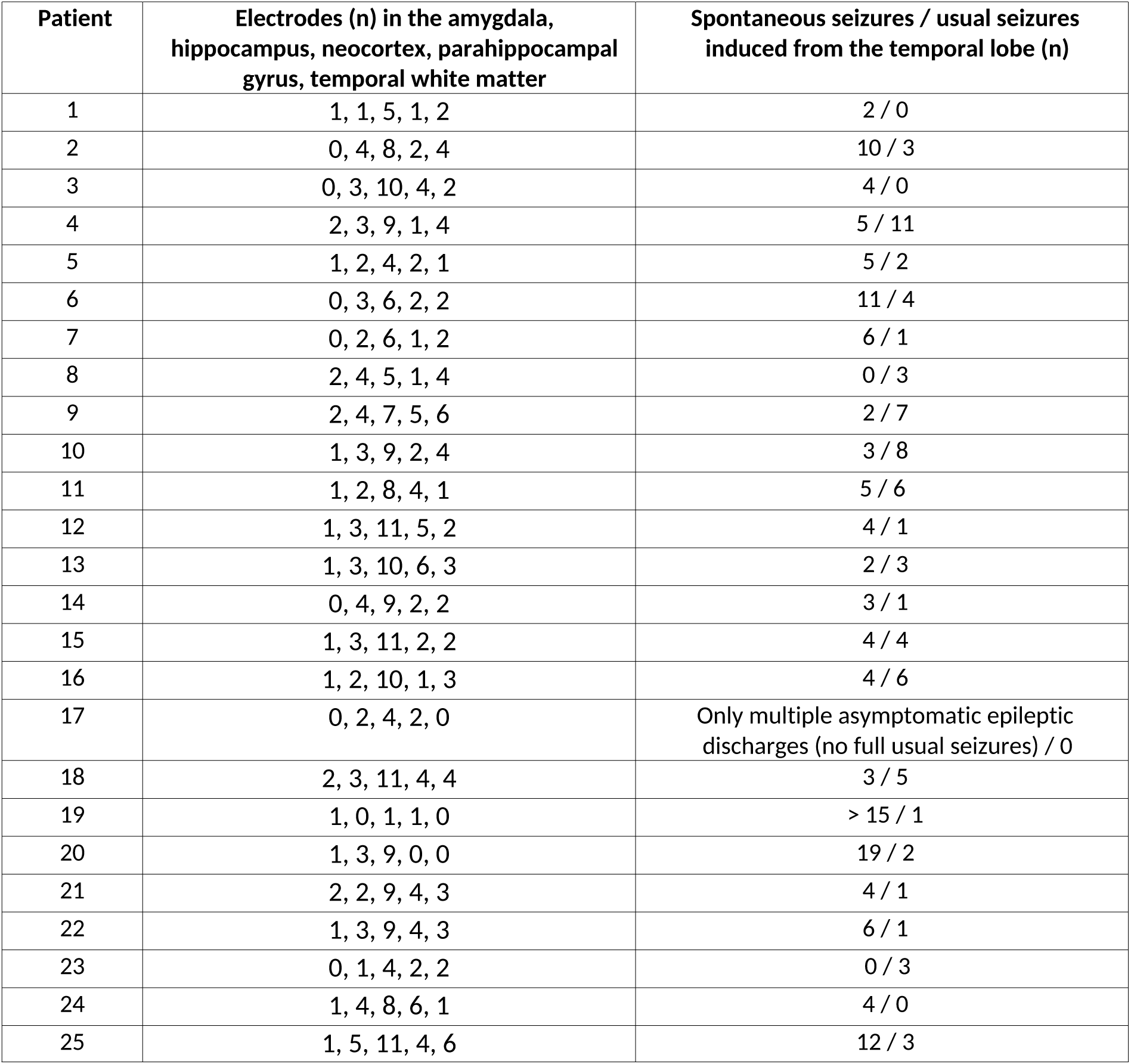
Supplementary clinical characteristics.

| Patient | Electrodes (n) in the amygdala, hippocampus, neocortex, parahippocampal gyrus, temporal white matter | Spontaneous seizures / usual seizures induced from the temporal lobe (n) |
| --- | --- | --- |
| 1 | 1, 1, 5, 1, 2 | 2 / 0 |
| 2 | 0, 4, 8, 2, 4 | 10 / 3 |
| 3 | 0, 3, 10, 4, 2 | 4 / 0 |
| 4 | 2, 3, 9, 1, 4 | 5 / 11 |
| 5 | 1, 2, 4, 2, 1 | 5 / 2 |
| 6 | 0, 3, 6, 2, 2 | 11 / 4 |
| 7 | 0, 2, 6, 1, 2 | 6 / 1 |
| 8 | 2, 4, 5, 1, 4 | 0 / 3 |
| 9 | 2, 4, 7, 5, 6 | 2 / 7 |
| 10 | 1, 3, 9, 2, 4 | 3 / 8 |
| 11 | 1, 2, 8, 4, 1 | 5 / 6 |
| 12 | 1, 3, 11, 5, 2 | 4 / 1 |
| 13 | 1, 3, 10, 6, 3 | 2 / 3 |
| 14 | 0, 4, 9, 2, 2 | 3 / 1 |
| 15 | 1, 3, 11, 2, 2 | 4 / 4 |
| 16 | 1, 2, 10, 1, 3 | 4 / 6 |
| 17 | 0, 2, 4, 2, 0 | Only multiple asymptomatic epileptic discharges (no full usual seizures) / 0 |
| 18 | 2, 3, 11, 4, 4 | 3 / 5 |
| 19 | 1, 0, 1, 1, 0 | > 15 / 1 |
| 20 | 1, 3, 9, 0, 0 | 19 / 2 |
| 21 | 2, 2, 9, 4, 3 | 4 / 1 |
| 22 | 1, 3, 9, 4, 3 | 6 / 1 |
| 23 | 0, 1, 4, 2, 2 | 0 / 3 |
| 24 | 1, 4, 8, 6, 1 | 4 / 0 |
| 25 | 1, 5, 11, 4, 6 | 12 / 3 |

**Supplementary Table S2:**
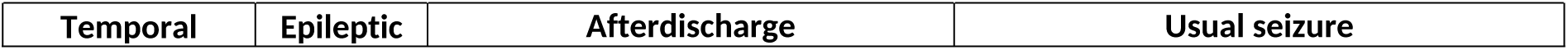
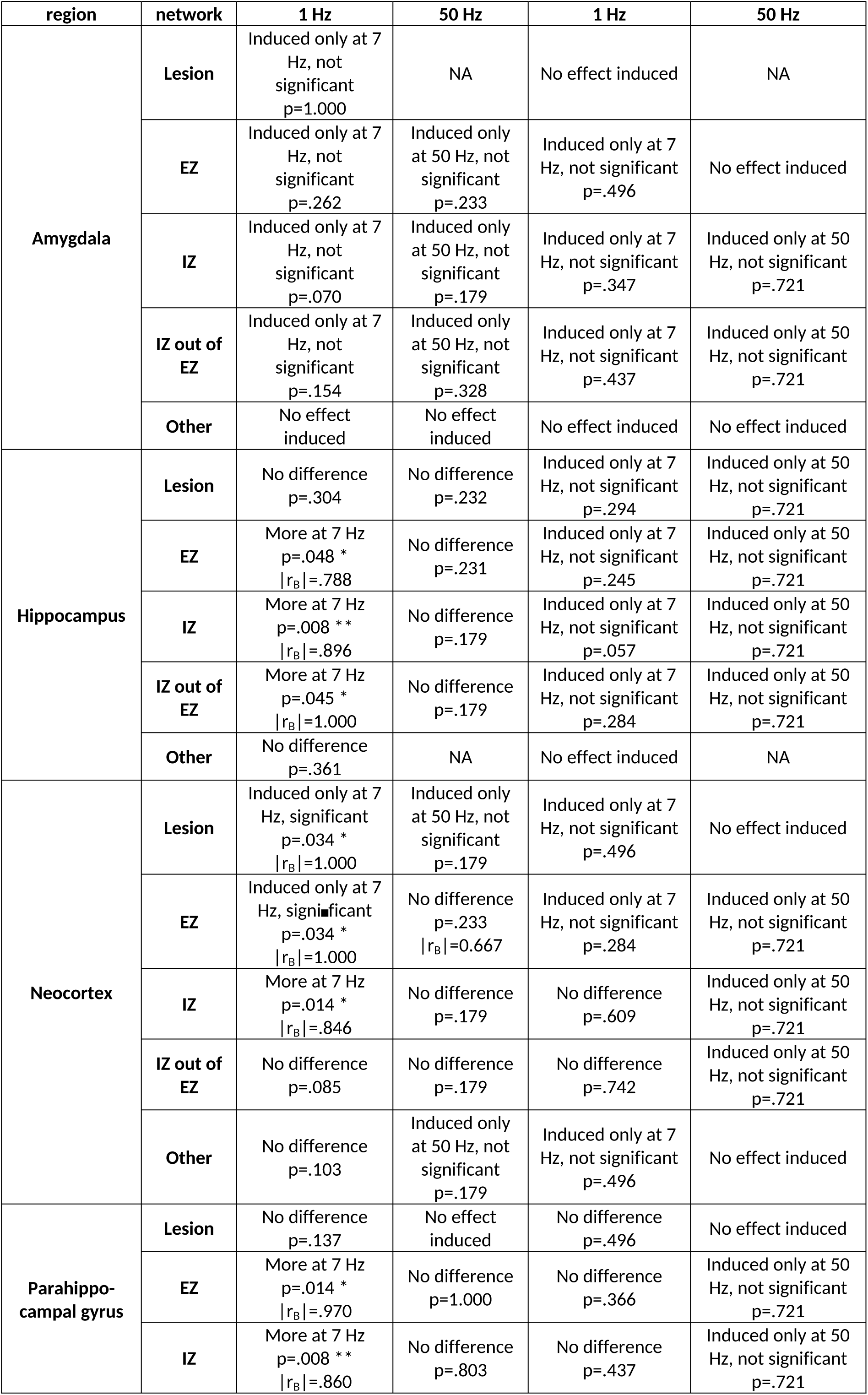
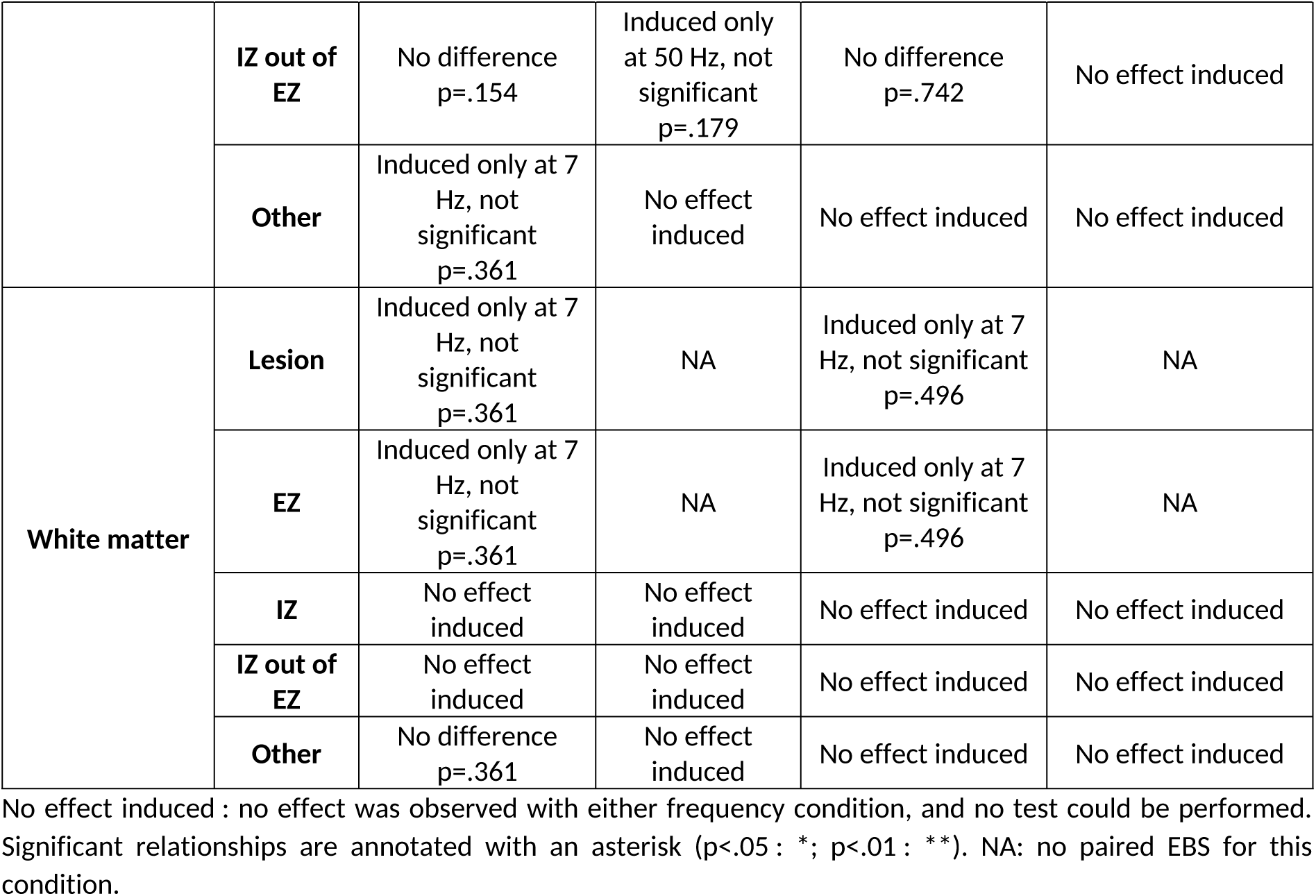
Statistical results (FDR-corrected p-value and effect size when significant relationship) for afterdischarge or usual seizure occurrence according to temporal structure and location in the epileptic network, for comparisons of 7-Hz EBS to 1-Hz or to 50-Hz EBS, at identical intensity and duration.

| region | network | 1 Hz | 50 Hz | 1 Hz | 50 Hz |
| --- | --- | --- | --- | --- | --- |
| Amygdala | Lesion | Induced only at 7 Hz, not significant<br>p=1.000 | NA | No effect induced | NA |
|  | EZ | Induced only at 7 Hz, not significant<br>p=.262 | Induced only at 50 Hz, not significant<br>p=.233 | Induced only at 7 Hz, not significant<br>p=.496 | No effect induced |
|  | IZ | Induced only at 7 Hz, not significant<br>p=.070 | Induced only at 50 Hz, not significant<br>p=.179 | Induced only at 7 Hz, not significant<br>p=.347 | Induced only at 50 Hz, not significant<br>p=.721 |
|  | IZ out of EZ | Induced only at 7 Hz, not significant<br>p=.154 | Induced only at 50 Hz, not significant<br>p=.328 | Induced only at 7 Hz, not significant<br>p=.437 | Induced only at 50 Hz, not significant<br>p=.721 |
|  | Other | No effect induced | No effect induced | No effect induced | No effect induced |
| Hippocampus | Lesion | No difference<br>p=.304 | No difference<br>p=.232 | Induced only at 7 Hz, not significant<br>p=.294 | Induced only at 50 Hz, not significant<br>p=.721 |
|  | EZ | More at 7 Hz<br>p=.048 *<br>\|r<sub>B</sub>\|=.788 | No difference<br>p=.231 | Induced only at 7 Hz, not significant<br>p=.245 | Induced only at 50 Hz, not significant<br>p=.721 |
|  | IZ | More at 7 Hz<br>p=.008 **<br>\|r<sub>B</sub>\|=.896 | No difference<br>p=.179 | Induced only at 7 Hz, not significant<br>p=.057 | Induced only at 50 Hz, not significant<br>p=.721 |
|  | IZ out of EZ | More at 7 Hz<br>p=.045 *<br>\|r<sub>B</sub>\|=1.000 | No difference<br>p=.179 | Induced only at 7 Hz, not significant<br>p=.284 | Induced only at 50 Hz, not significant<br>p=.721 |
|  | Other | No difference<br>p=.361 | NA | No effect induced | NA |
| Neocortex | Lesion | Induced only at 7 Hz, significant<br>p=.034 *<br>\|r<sub>B</sub>\|=1.000 | Induced only at 50 Hz, not significant<br>p=.179 | Induced only at 7 Hz, not significant<br>p=.496 | No effect induced |
|  | EZ | Induced only at 7 Hz, significant<br>p=.034 *<br>\|r<sub>B</sub>\|=1.000 | No difference<br>p=.233<br>\|r<sub>B</sub>\|=0.667 | Induced only at 7 Hz, not significant<br>p=.284 | Induced only at 50 Hz, not significant<br>p=.721 |
|  | IZ | More at 7 Hz<br>p=.014 *<br>\|r<sub>B</sub>\|=.846 | No difference<br>p=.179 | No difference<br>p=.609 | Induced only at 50 Hz, not significant<br>p=.721 |
|  | IZ out of EZ | No difference<br>p=.085 | No difference<br>p=.179 | No difference<br>p=.742 | Induced only at 50 Hz, not significant<br>p=.721 |
|  | Other | No difference<br>p=.103 | Induced only at 50 Hz, not significant<br>p=.179 | Induced only at 7 Hz, not significant<br>p=.496 | No effect induced |
| Parahippocampal gyrus | Lesion | No difference<br>p=.137 | No effect induced | No difference<br>p=.496 | No effect induced |
|  | EZ | More at 7 Hz<br>p=.014 *<br>\|r<sub>B</sub>\|=.970 | No difference<br>p=1.000 | No difference<br>p=.366 | Induced only at 50 Hz, not significant<br>p=.721 |
|  | IZ | More at 7 Hz<br>p=.008 **<br>\|r<sub>B</sub>\|=.860 | No difference<br>p=.803 | No difference<br>p=.437 | Induced only at 50 Hz, not significant<br>p=.721 |
|  | **IZ out of EZ** | No difference<br>p=.154 | Induced only<br>at 50 Hz, not<br>significant<br>p=.179 | No difference<br>p=.742 | No effect induced |
|  | **Other** | Induced only at 7<br>Hz, not<br>significant<br>p=.361 | No effect<br>induced | No effect induced | No effect induced |
| **White matter** | **Lesion** | Induced only at 7<br>Hz, not<br>significant<br>p=.361 | NA | Induced only at 7<br>Hz, not significant<br>p=.496 | NA |
|  | **EZ** | Induced only at 7<br>Hz, not<br>significant<br>p=.361 | NA | Induced only at 7<br>Hz, not significant<br>p=.496 | NA |
|  | **IZ** | No effect<br>induced | No effect<br>induced | No effect induced | No effect induced |
|  | **IZ out of EZ** | No effect<br>induced | No effect<br>induced | No effect induced | No effect induced |
|  | **Other** | No difference<br>p=.361 | No effect<br>induced | No effect induced | No effect induced |

No effect induced : no effect was observed with either frequency condition, and no test could be performed. Significant relationships are annotated with an asterisk (p<.05 : \*; p<.01 : \*\*). NA: no paired EBS for this condition.

**Supplementary Table S3:**
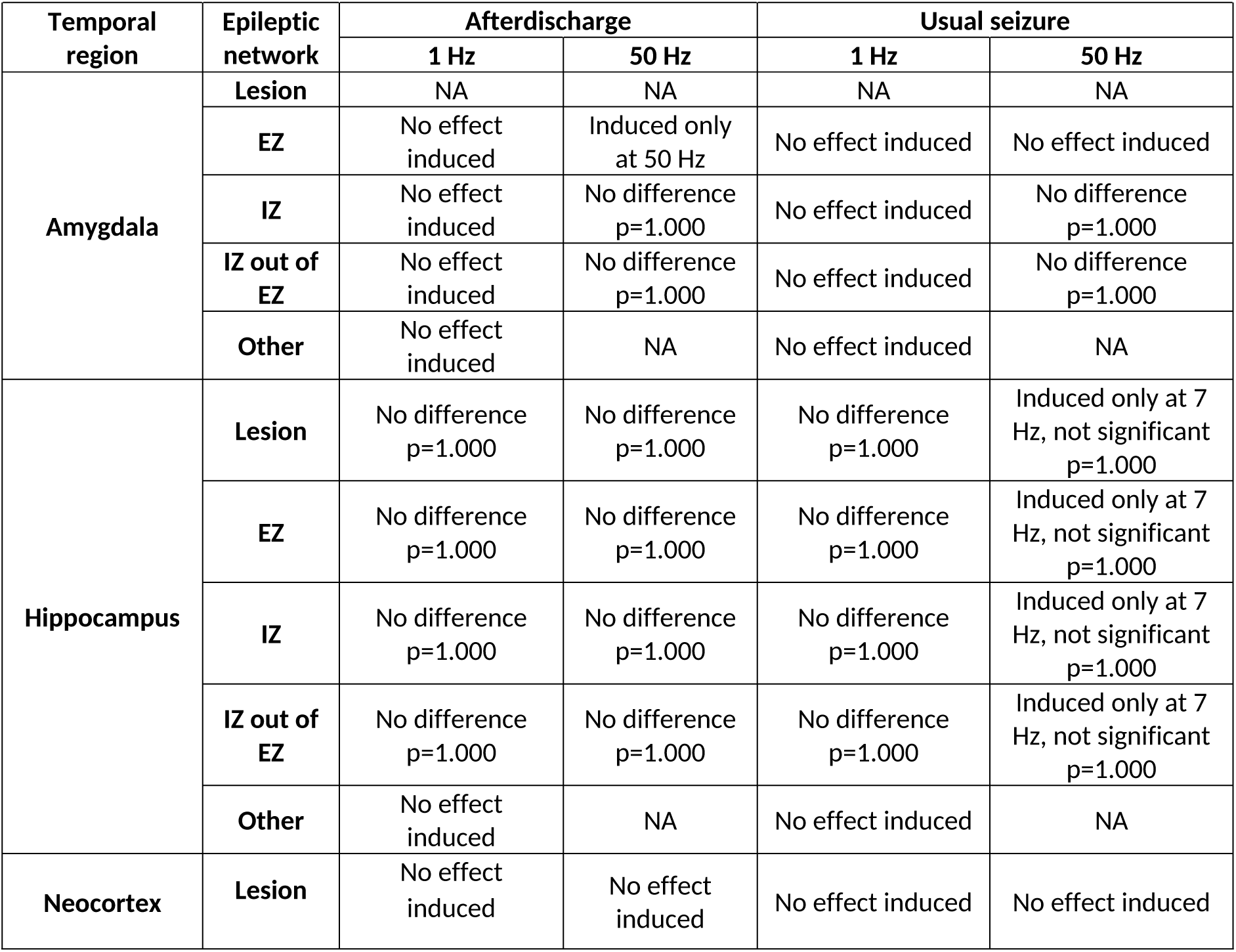
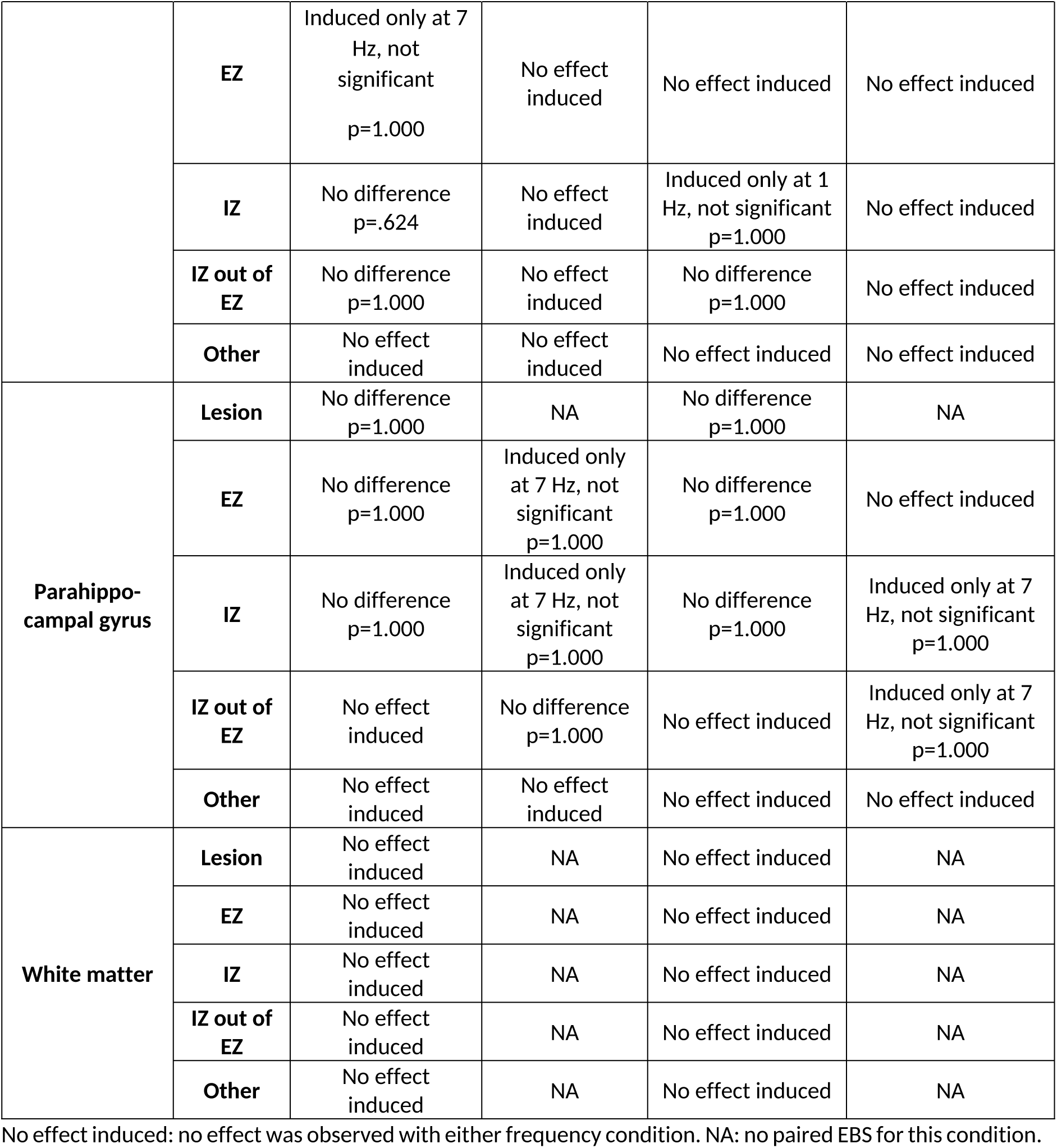
Statistical results (FDR-corrected p-value) for afterdischarge or usual seizure occurrence according to temporal structure and location in the epileptic network, for comparisons of 7-Hz EBS to 1-Hz or to 50-Hz EBS, at similar total charge quantity.

**Supplementary Table S4:**
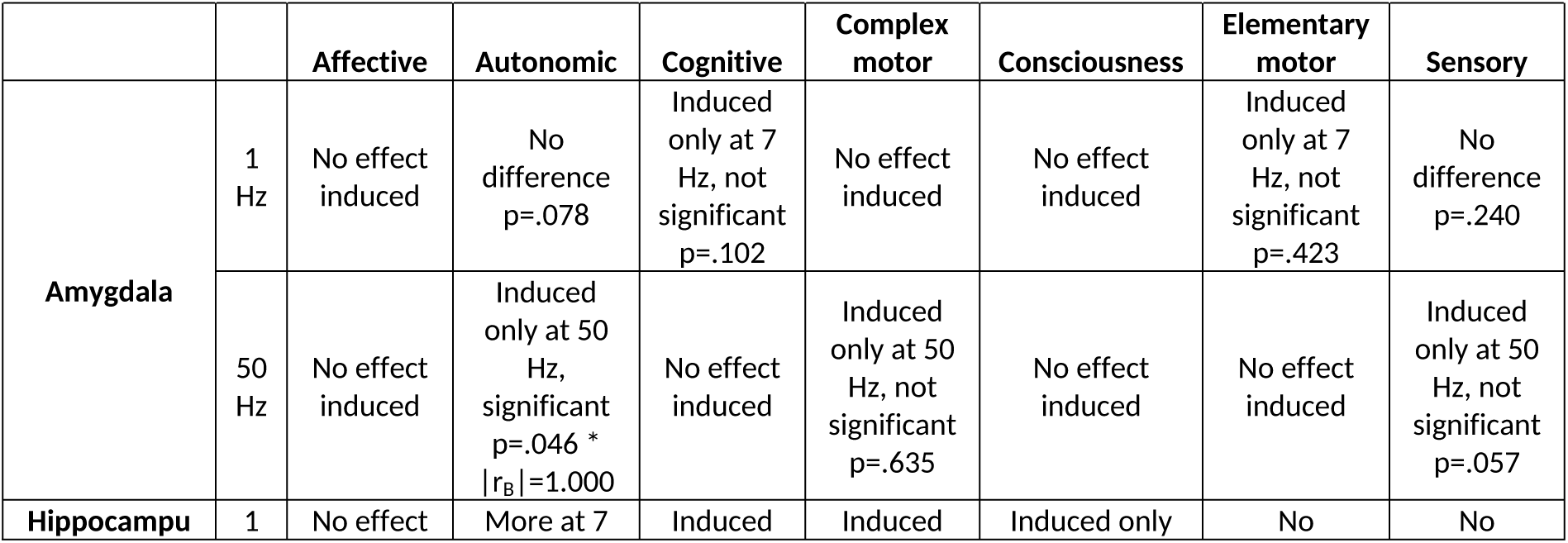
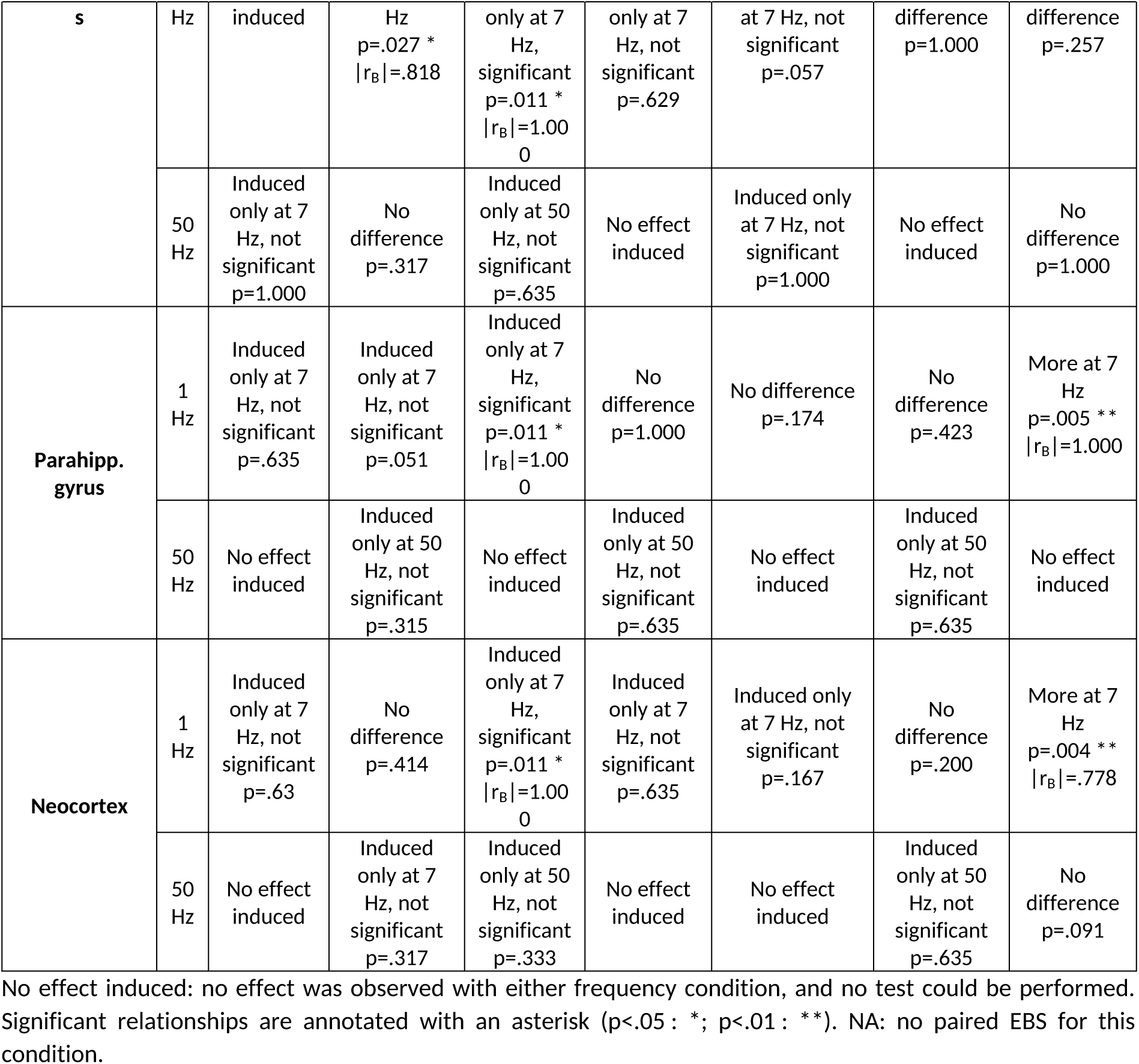
Statistical results (FDR-corrected p-value and effect size when significant relationship) for clinical sign occurrence according to temporal structure, for comparisons of 7-Hz EBS to 1-Hz or to 50-Hz EBS, at identical intensity and duration.

**Supplementary Table S5:** Statistical results (FDR-corrected p-value) for clinical sign occurrence according to temporal structure, for comparisons of 7-Hz EBS to 1-Hz or to 50-Hz EBS, at similar total charge quantity.

|  |  | **Affective** | **Autonomic** | **Cognitive** | **Complex motor** | **Consciousness** | **Elementary motor** | **Sensory** |
| --- | --- | --- | --- | --- | --- | --- | --- | --- |
| **Amygdala** | **1 Hz** | No effect induced | No effect induced | No effect induced | No effect induced | No effect induced | No effect induced | Induced only at 7 Hz, not significant p=.715 |
|  | **50 Hz** | No effect induced | No difference p=.423 | Induced only at 50 Hz, not significant p=.423 | No effect induced | No effect induced | Induced only at 7 Hz, not significant p=.635 | Induced only at 50 Hz, not significant p=.317 |
| **Hippocampus** | **1 Hz** | Induced only at 7 Hz, not significant p=1.000 | No difference p=.791 | No difference p=.873 | Induced only at 1 Hz, not significant p=.719 | No difference p=.359 | No effect induced | No difference p=.752 |
|  | 50 Hz | No effect induced | Induced only at 7 Hz, not significant p=.423 | Induced only at 7 Hz, not significant p=.423 | No effect induced | Induced only at 7 Hz, not significant p=1.000 | No effect induced | Induced only at 7 Hz, not significant p=.317 |
| Parahipp. gyrus | 1 Hz | No effect induced | No difference p=.570 | No difference p=1.000 | No effect induced | Induced only at 1 Hz, not significant p=.359 | No difference p=1.000 | No difference p=.715 |
|  | 50 Hz | No effect induced | No effect induced | No difference p=.873 | Induced only at 7 Hz, not significant p=1.000 | No difference p=1.000 | Induced only at 7 Hz p=0.635 | Induced only at 7 Hz, not significant p=.317 |
| Neocortex | 1 Hz | No effect induced | Induced only at 7 Hz, not significant p=.570 | Induced only at 7 Hz, not significant p=.873 | No effect induced | No effect induced | No effect induced | No difference p=.752 |
|  | 50 Hz | No effect induced | Induced only at 7 Hz, not significant p=.423 | No effect induced | No effect induced | No effect induced | No effect induced | Induced only at 50 Hz, not significant p=.317 |

No effect induced: no effect was observed with either frequency condition. NA: no EBS for this condition.

**Supplementary Table S6:**
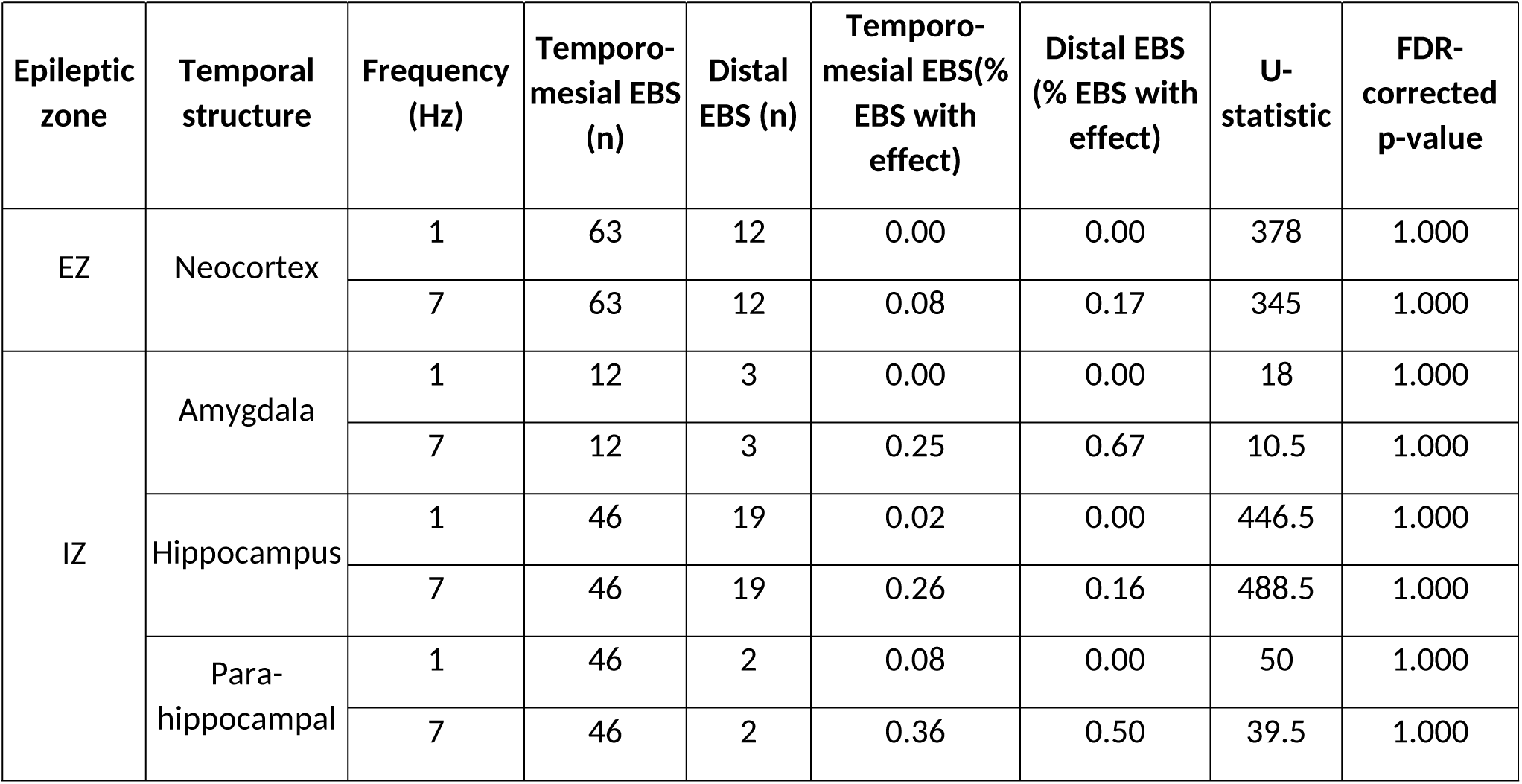
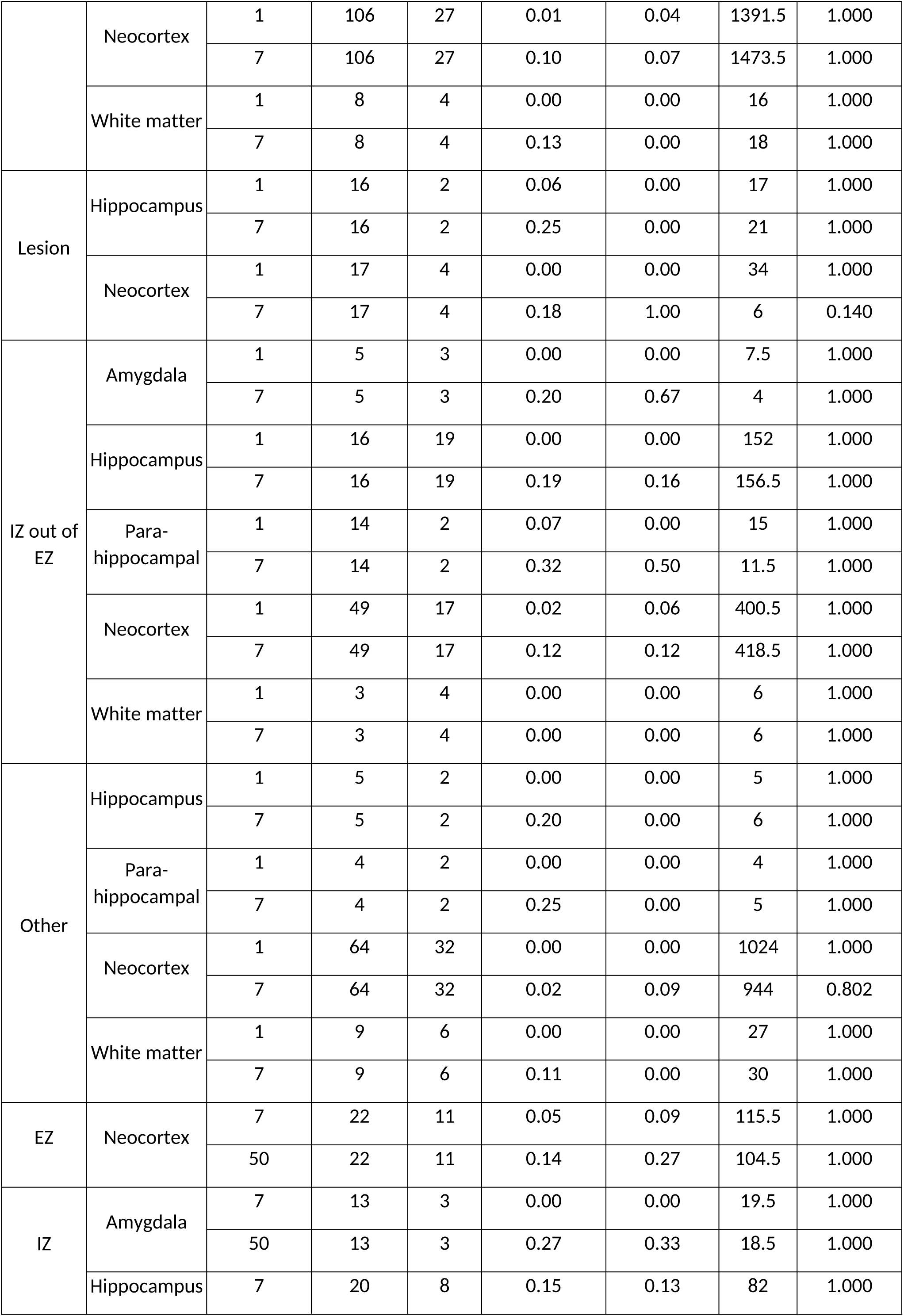
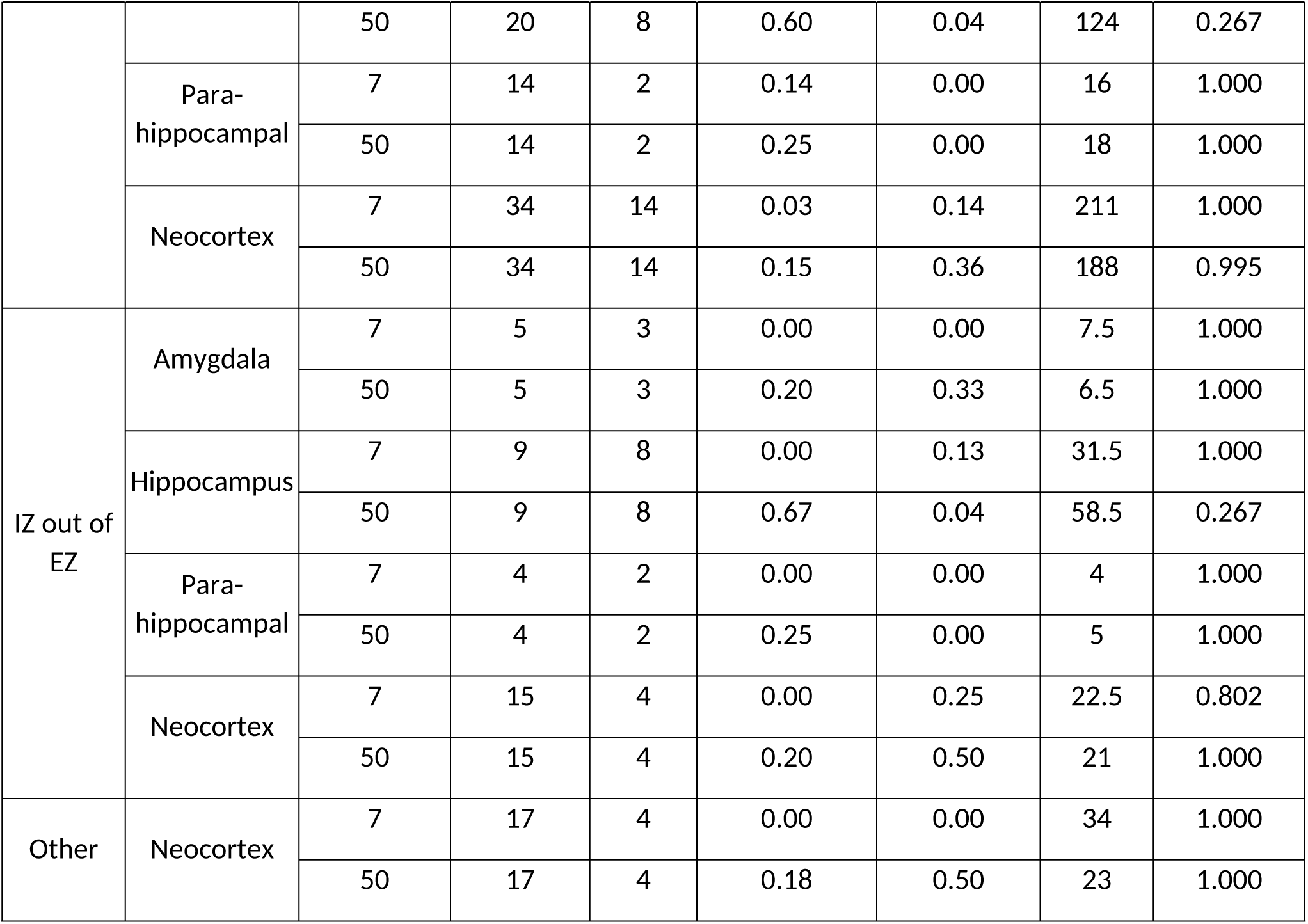
Statistical results of comparisons between patients with temporo-mesial and non-temporo-mesial EZ for afterdischarge occurrence, stratified by temporal structure and epileptic zone. Comparisons were led for each frequency, in 7-Hz versus 1-Hz comparisons and in 7-Hz versus 50-Hz comparisons delivered at identical intensity and duration. Comparisons are reported only for conditions in which at least one EBS was available in each patient group.

**Supplementary Table S7:**
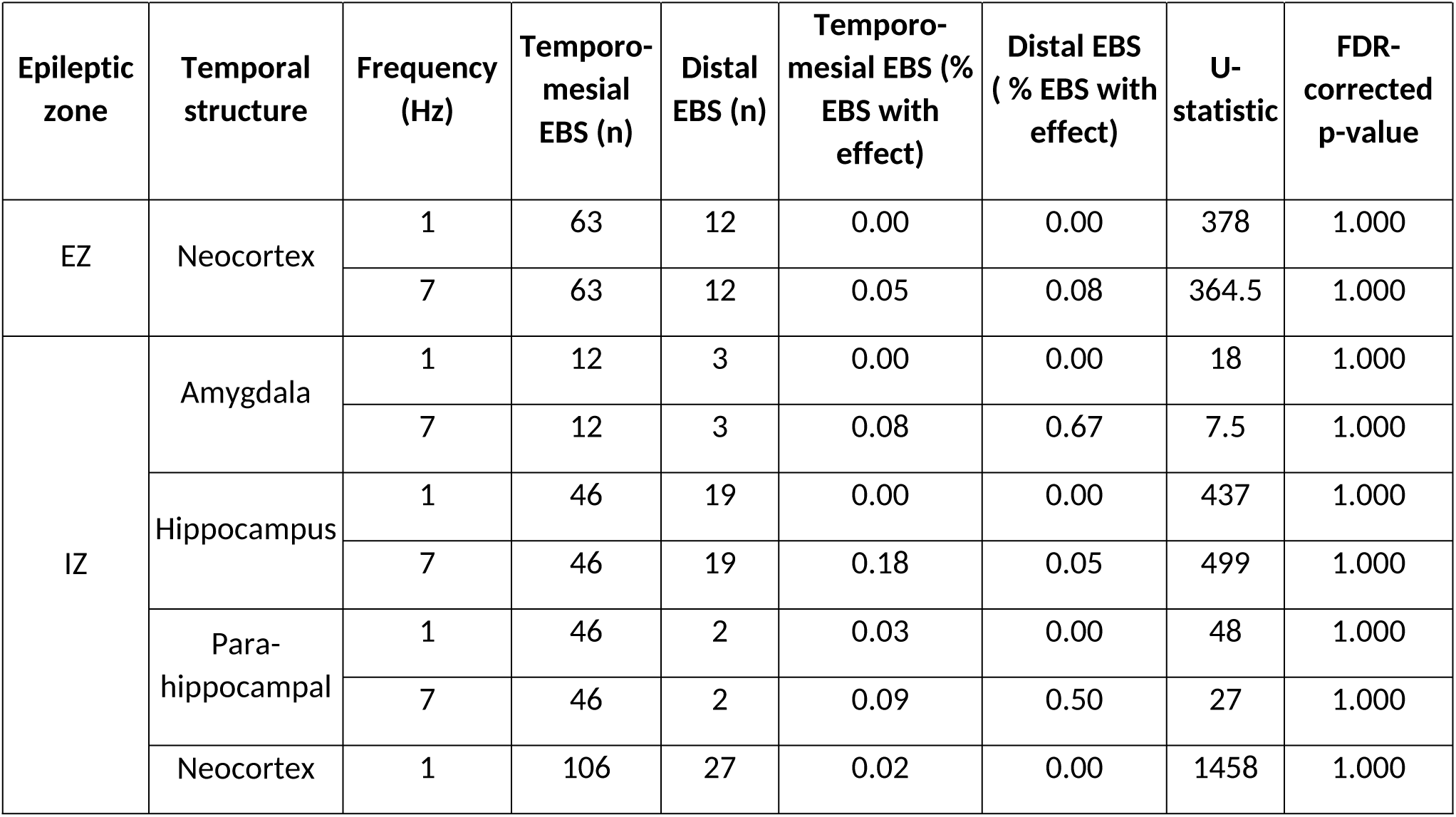
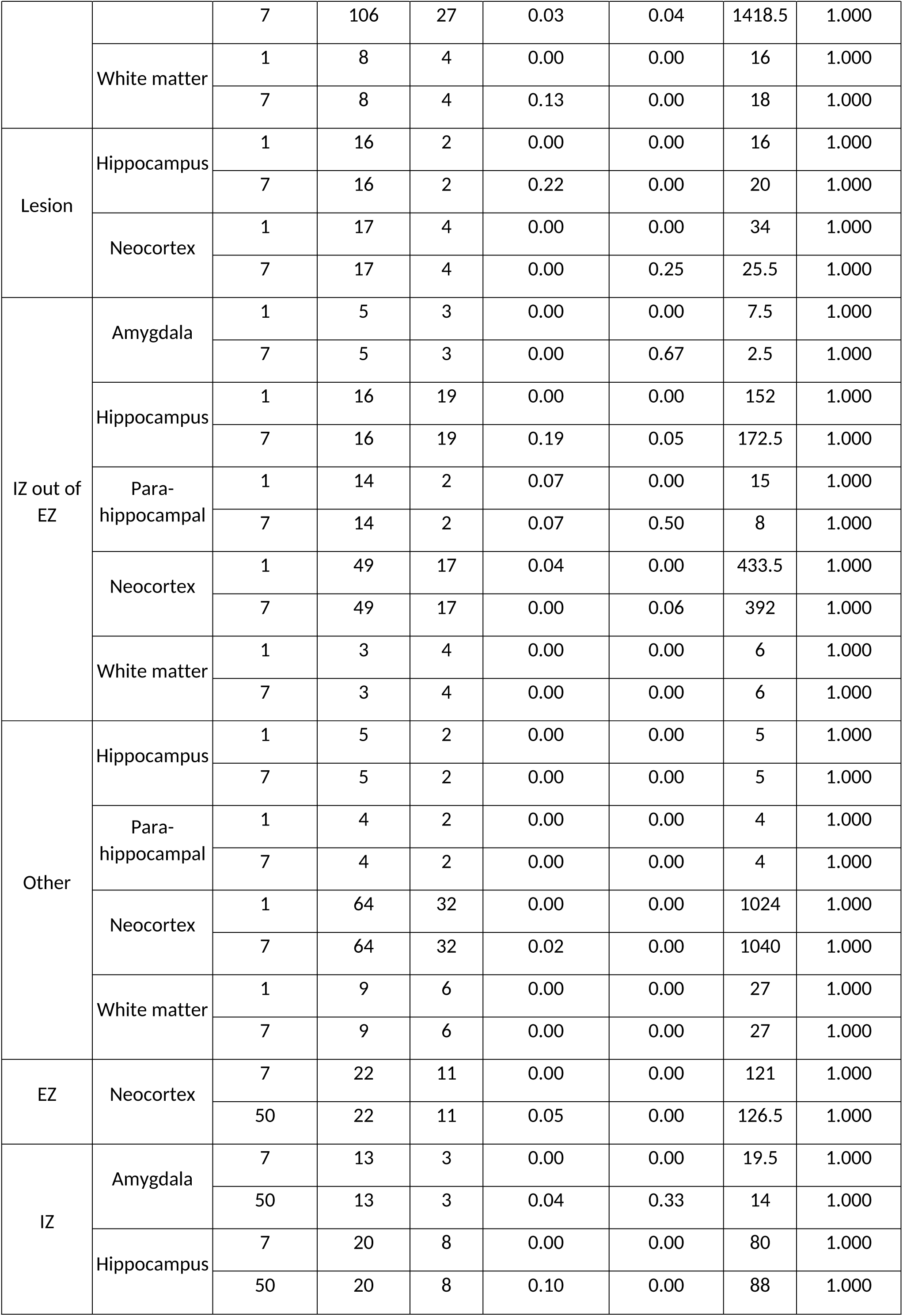
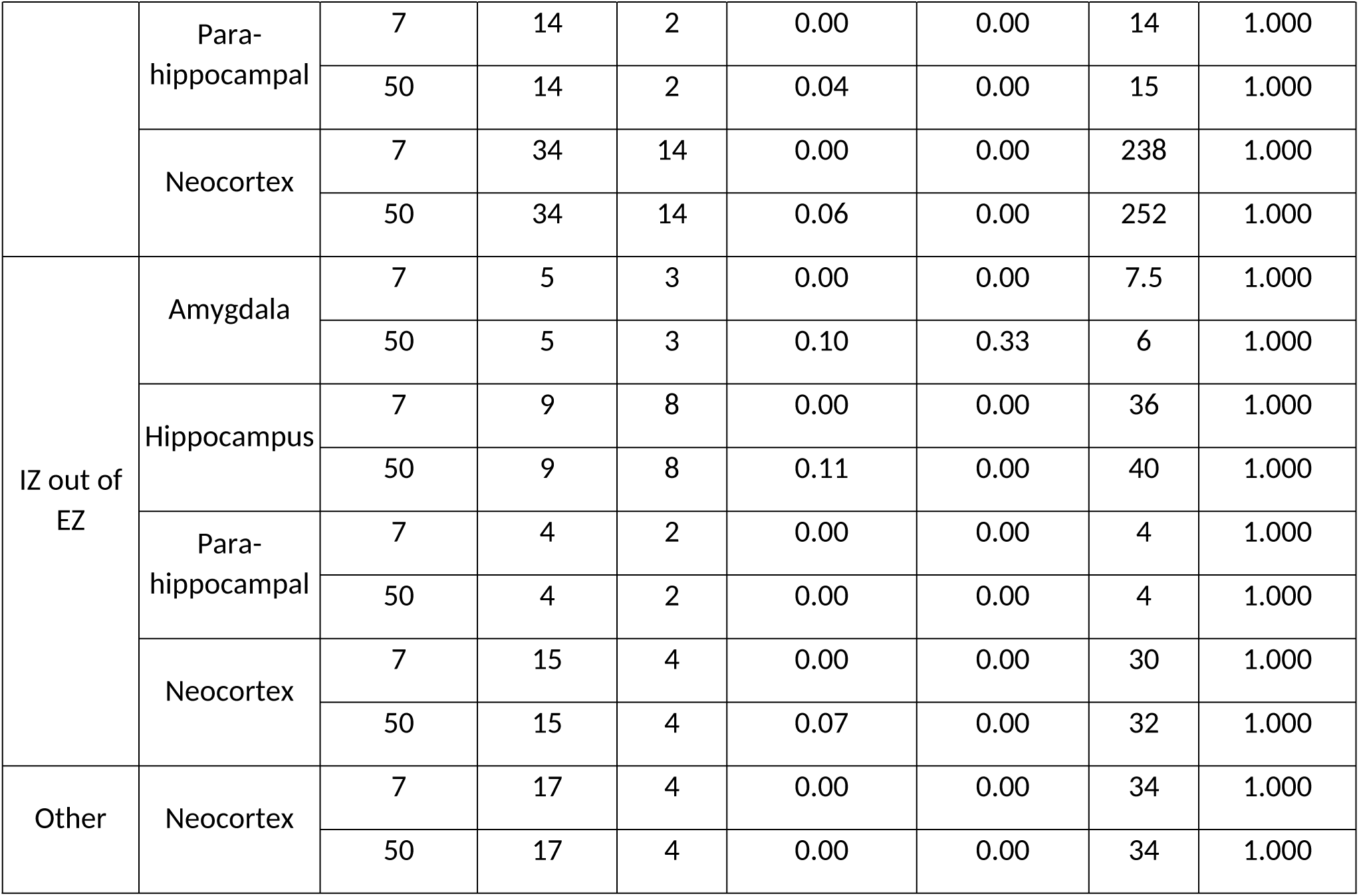
Statistical results of comparisons between patients with temporo-mesial and non-temporo-mesial EZ for usual seizure occurrence, stratified by temporal structure and epileptic zone. Comparisons were led for each frequency, in 7-Hz versus 1-Hz comparisons and in 7-Hz versus 50-Hz comparisons delivered at identical intensity and duration. Comparisons are reported only for conditions in which at least one EBS was available in each patient group.

**Supplementary Table S8:**
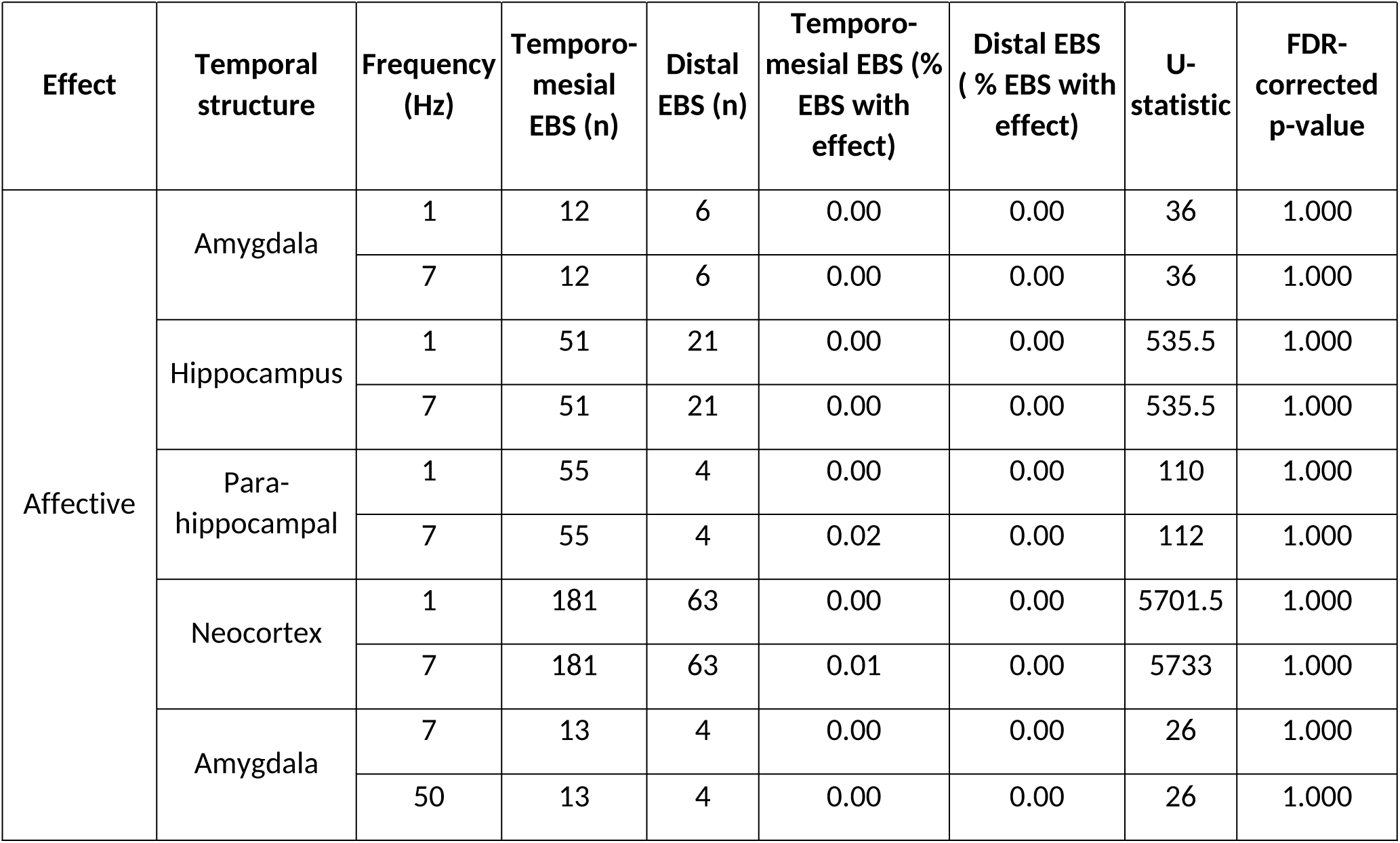
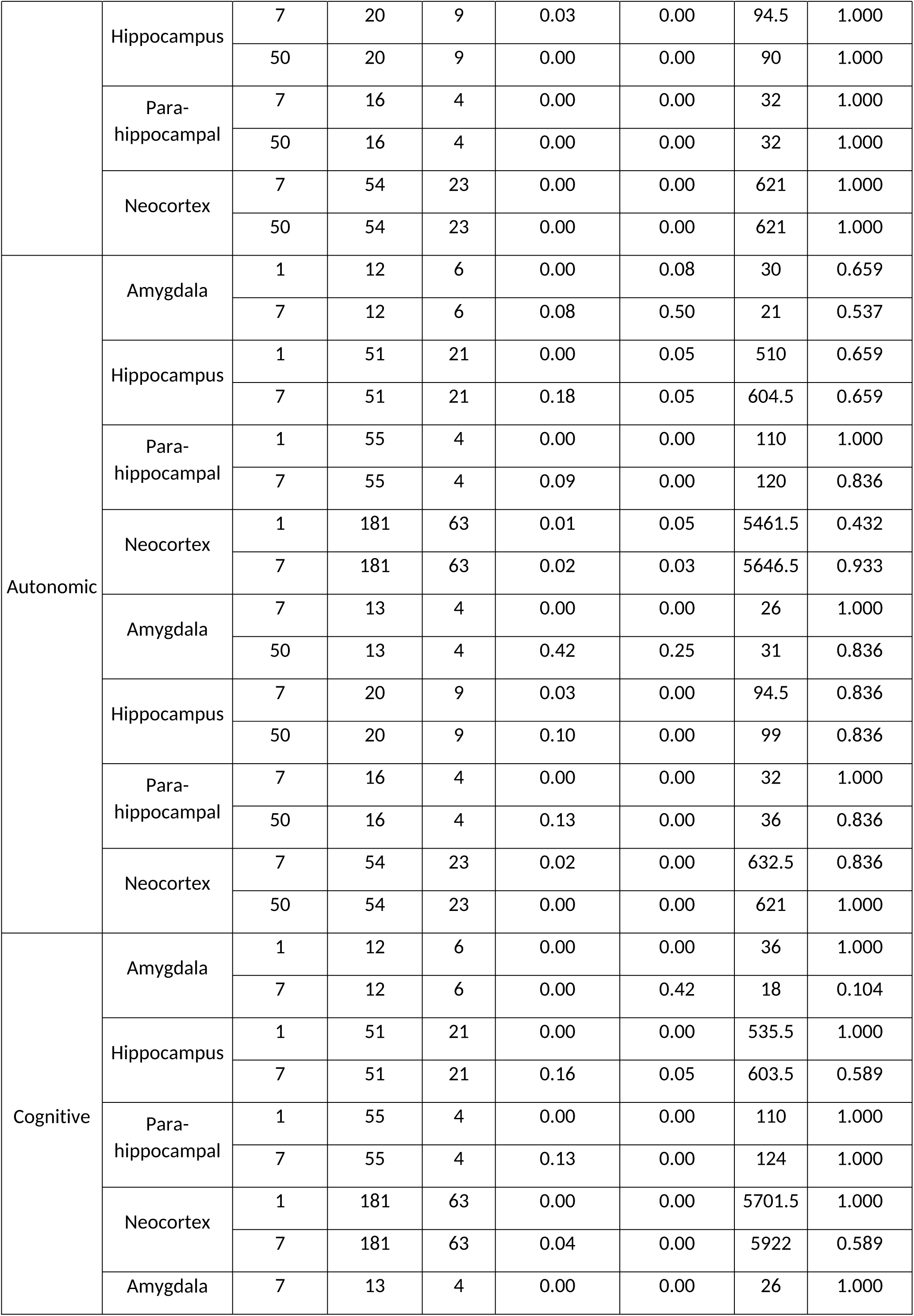
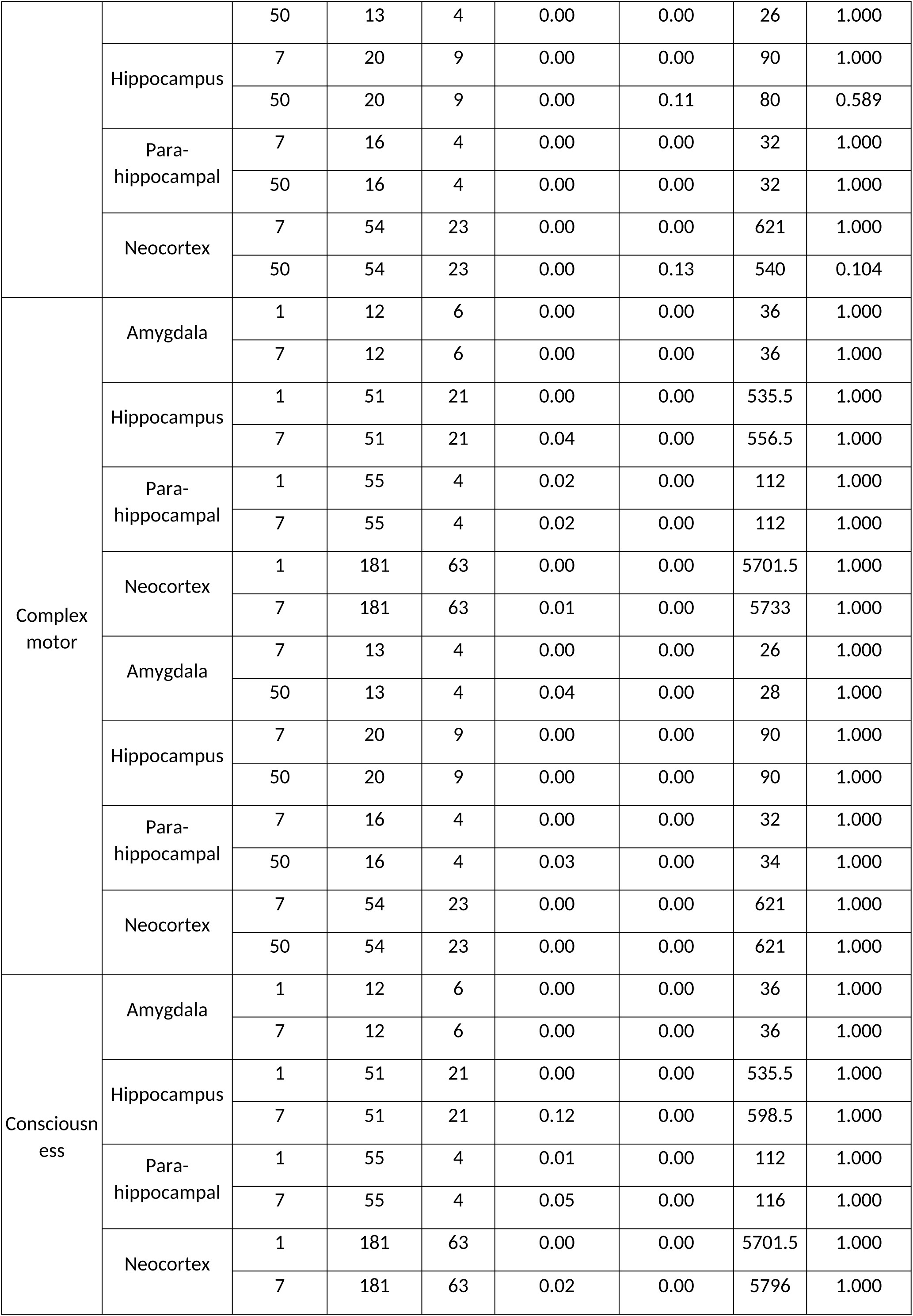
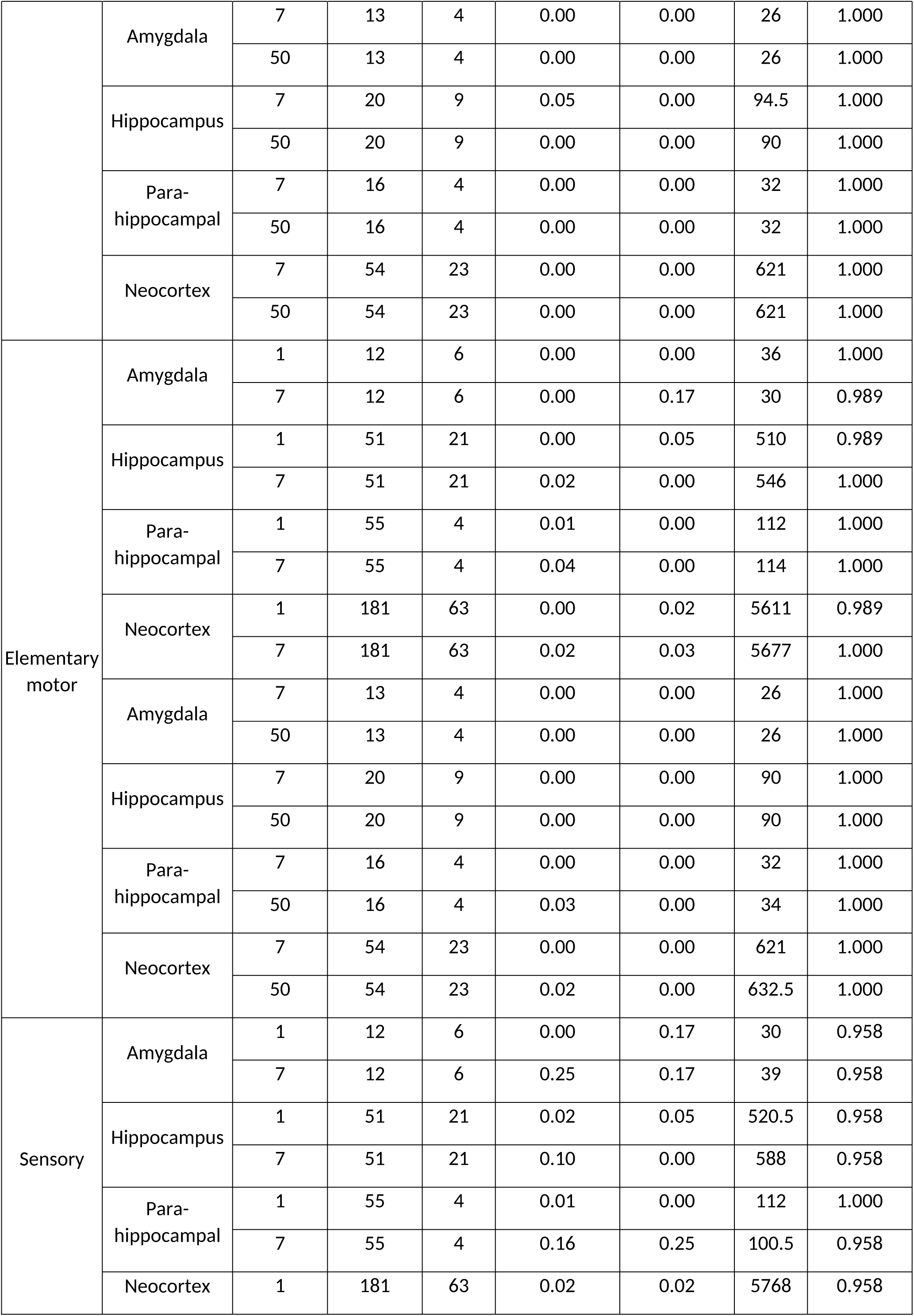
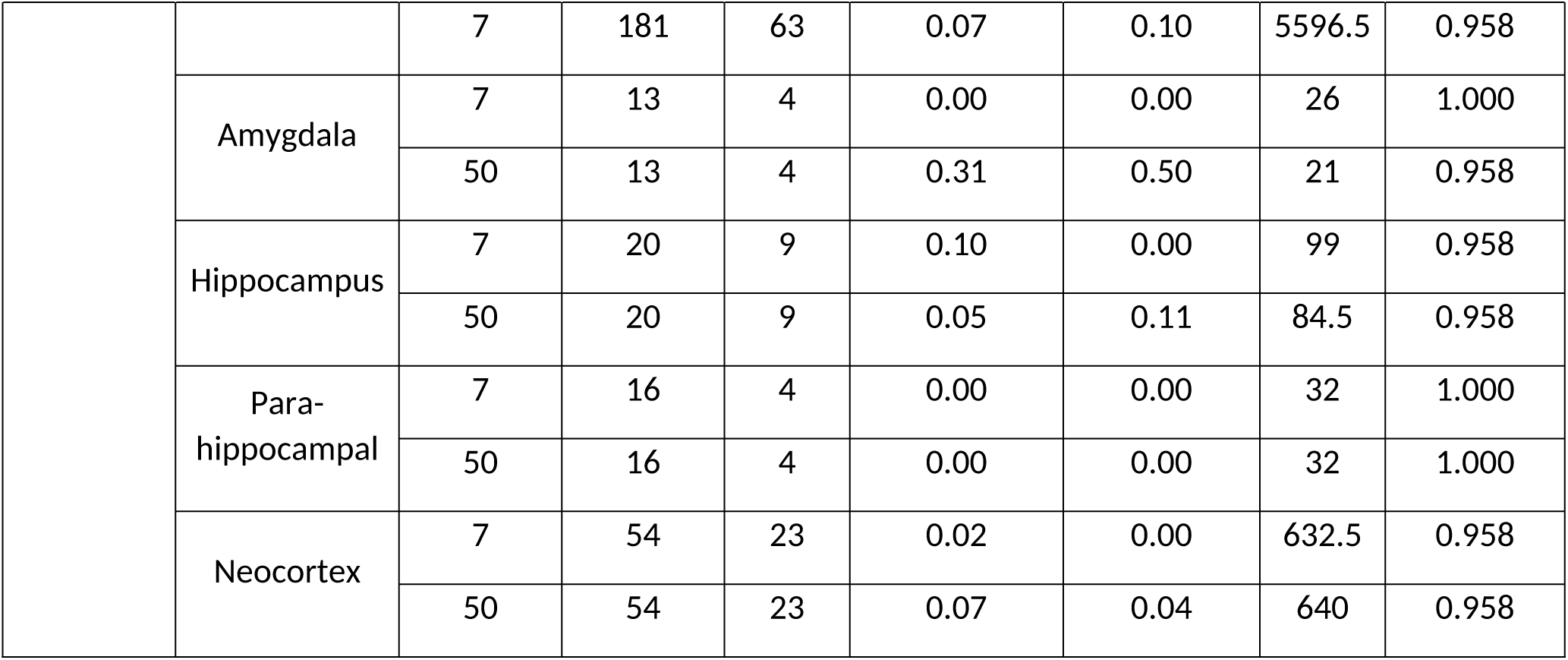
Statistical results of comparisons between patients with temporo-mesial and non-temporo-mesial EZ for clinical sign occurrence, stratified by temporal structure and epileptic zone. Comparisons were led for each frequency, in 7-Hz versus 1-Hz comparisons and in 7-Hz versus 50-Hz comparisons delivered at identical intensity and duration. Comparisons are reported only for conditions in which at least one EBS was available in each patient group.

## Notes

### Competing Interest Statement

The authors have declared no competing interest.

### Clinical Trial

NCT03738072

### Funding Statement

the French National Research Agency as part of the "DYNEUMICS" project, reference ANR-21-CE17-0029, and by the Toulouse University Hospital (local grant: ARI StiMiC, NCT03738072)

### Author Declarations

Ethics Committee (CPP Ile de France VII; ID:18-026), and the French National Agency for Medicines and Health Products Safety (ANSM; 2018-A00383-52)

