## Supplementary figures and images for "Theta-Range SEEG Stimulation for Temporal Lobe Mapping: An Alternative to Conventional 1-Hz and 50-Hz Protocols"

### Supplementary Figure 1

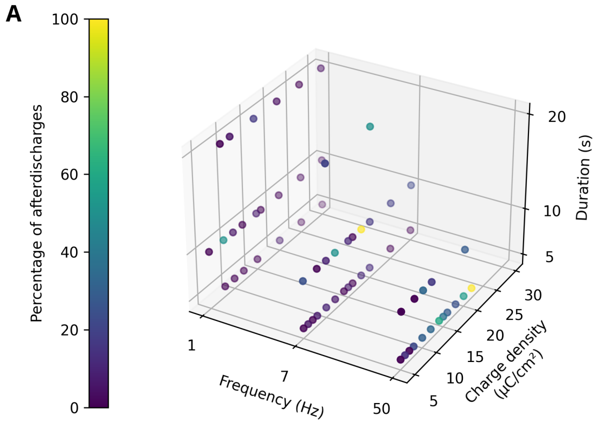

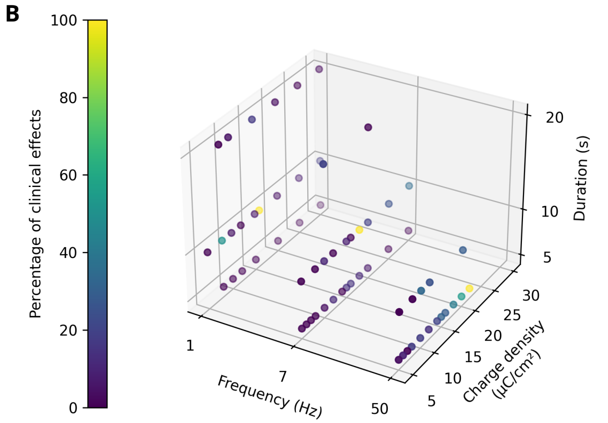

### Supplementary Figure 2

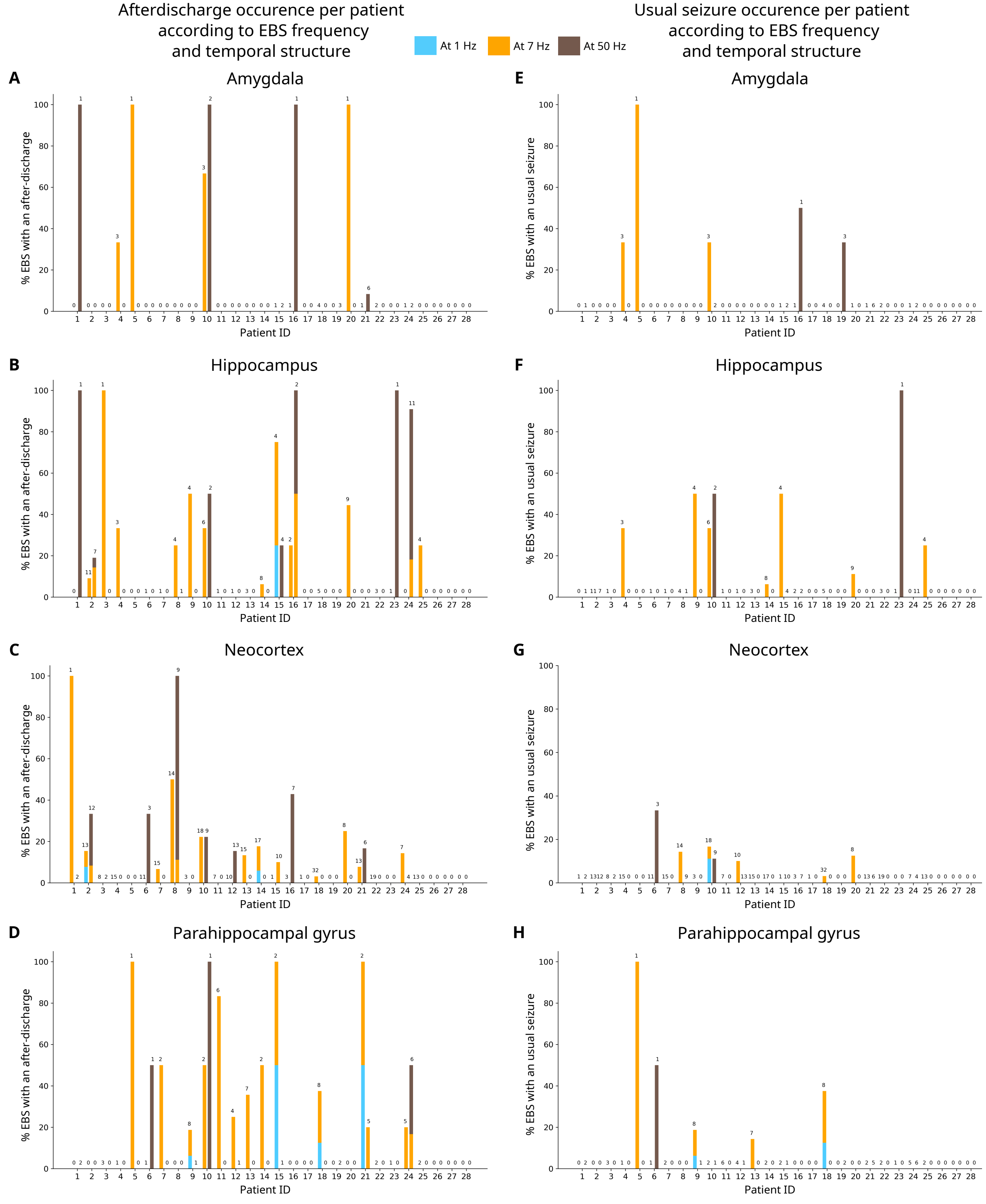

### Supplementary Figure 3

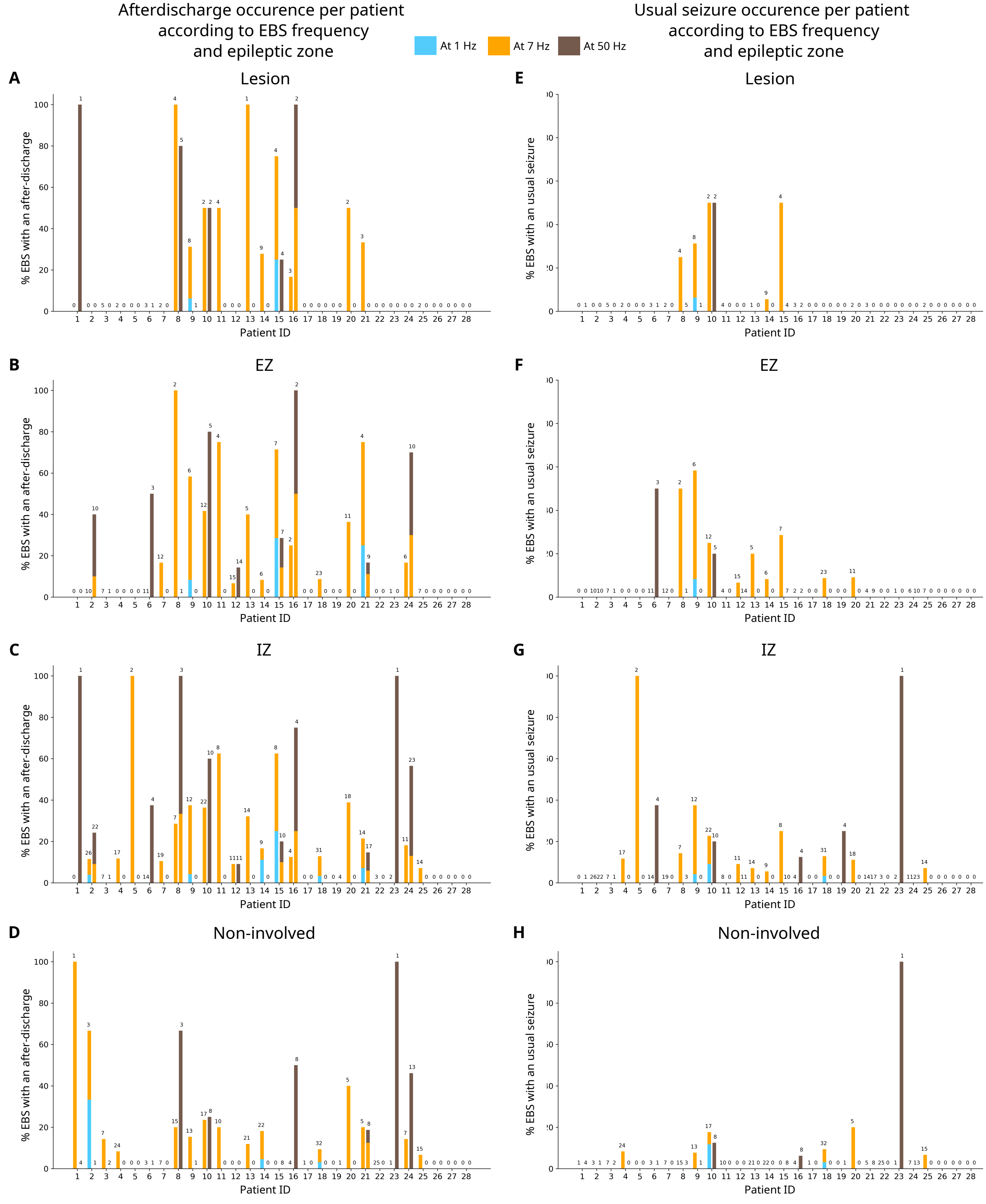

### Supplementary Figure 4

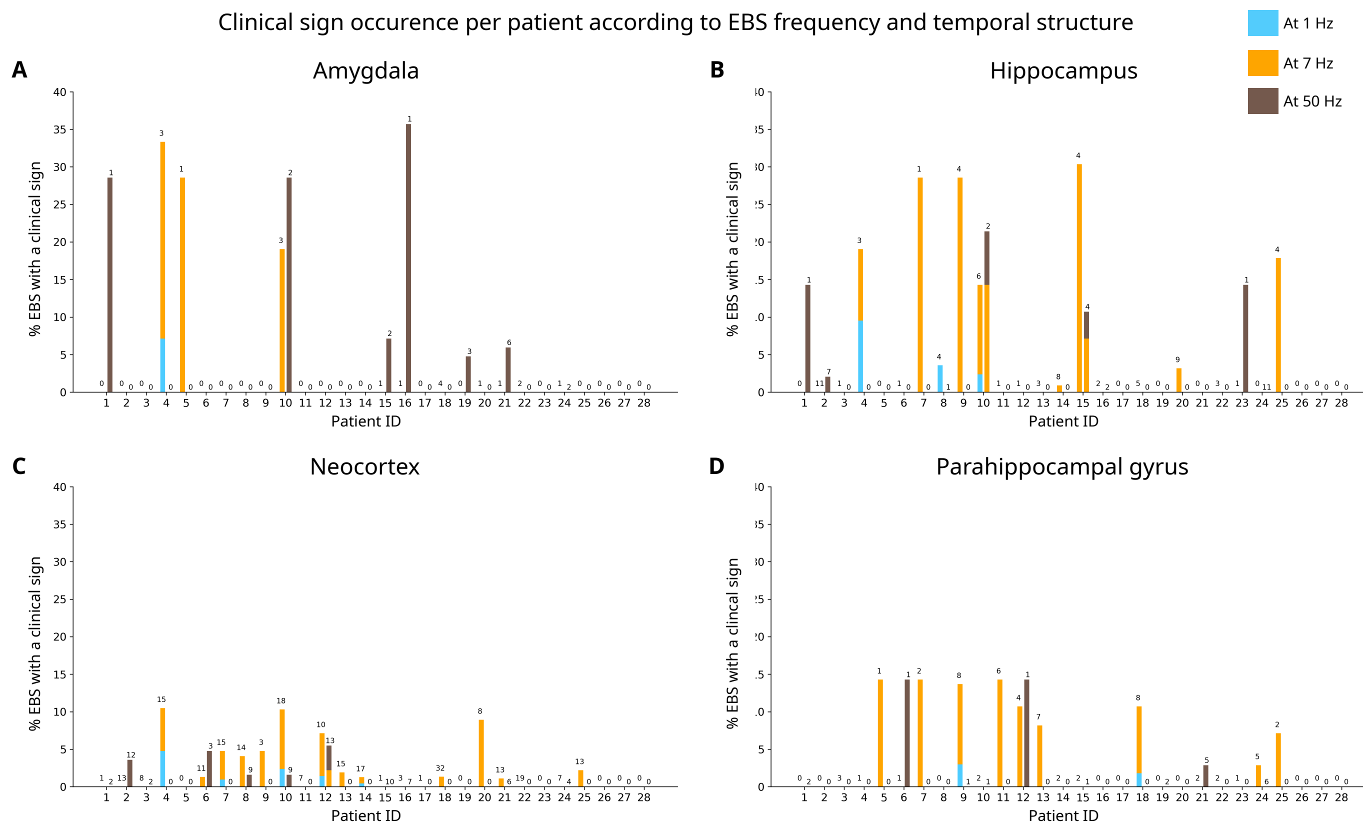
